# Large-scale assessment of socioeconomic, demographic and health system structures with US county excess mortality, 2020-2024

**DOI:** 10.64898/2026.07.04.26357291

**Authors:** Michael Levitt, Ben Marten, Gal Oren, John P.A. Ioannidis

**Affiliations:** Department of Structural Biology, Stanford University School of Medicine, Stanford, California, USA; Mortality.Watch, St. Petersburg, Florida, USA; Computer Science Department, Technion Israel Institute of Technology, Haifa, Israel; Meta-Research Innovation Center at Stanford (METRICS), Stanford University School of Medicine, Stanford, California, USA; Stanford Prevention Research Center, Department of Medicine, Stanford University School of Medicine, Stanford, California, USA; Department of Epidemiology and Population Health, Stanford University School of Medicine, Stanford, California, USA; Department of Biomedical Data Science, Stanford University School of Medicine, Stanford, California, USA

**Keywords:** COVID-19, excess mortality, US counties, social determinants, Area Health Resources File, dimensionality

## Abstract

Socioeconomic, demographic and health system structures may have shaped COVID-19 impact, but past analyses examined few factors. We systematically examined correlates of COVID-era excess mortality, considering 2,745 county-level variables of demography, race/ethnicity, income, insurance, education, employment, housing, and health system. Pearson correlation coefficients (CCs) were obtained for the most recent available pre-pandemic value against population-weighted age-standardized county excess-death for each year during 2020–2024. Variables were grouped by meaning into 11 semantic super-clusters. Overall, 24.1% of variables reached at least a moderate correlation level (|CC| > 0.30) and 7.4% reached strong correlations (| CC| > 0.45). Strongest correlations were seen for poverty share (CC +0.64), college attainment (−0.62), and high income (−0.61). At least moderate correlations were seen for 11.4% of variables in 2020 and 14.8% in 2021, but only 3.3%, 1.3%, and 4.1% in 2022, 2023, and 2024, respectively. The concentration in 2020–2021 was sharpest in the non-elderly population. Of 656 variables with |CC| > 0.30, 295/454 moderate-band and 200/202 strong-band variables belonged to demography and socioeconomic super-clusters. Only 10% of health system variables reached | CC| > 0.30, versus 42% of socioeconomic and demographic variables. Using the most recent available value until 2023 or 2015, different population weighting, and Spearman correlations left the direction and the leading variables unchanged, but the correlation magnitudes scale with the weight given to large counties. Overall, these ecological analyses suggest stronger relationships of socioeconomic structure and demographics than of baseline health-system structure and supply with excess mortality, especially during 2020–2021.

## INTRODUCTION

The COVID-19 pandemic demonstrated large geographical variability in its footprint and revealed the high vulnerability of populations with disadvantaged status. County-level analyses of COVID-19 mortality have repeatedly implicated poverty, race, lack of insurance, and poor health-care access as correlates of adverse outcome (1–8), but the literature is fragmented. Each study has typically picked only a handful of predictors; moreover, conclusions can hinge on what analyses are performed and reported.

There is increasing interest in studying epidemiological exposures comprehensively at large scale, extending the omics paradigm that is dominant in genetics to diverse types of social, environmental, and other exposures (9–11). Moreover, increasing attention is given to robustness analyses, where large-scale assessments can be examined for their sensitivity to analytical choices (12, 13). Here, we combined these two principles to assess the footprint of diverse county-level features on COVID-19 excess deaths across US counties in 2020–2024.

Instead of pre-selecting predictors, we screened an exhaustive set of 2,745 county-level variables. Moreover, given that COVID-19 epidemiological estimates are known to be sensitive to the underlying data and modeling assumptions (14), we examined the robustness of the results considering different sets of variables collected until just prior to the pandemic (2019), during the pandemic, or 5 or more years before the pandemic; different population weighting; and different correlation metrics. Our analyses aimed to identify clusters and super-clusters of variables; examine how many variables have at least moderate correlations with excess deaths; and characterize whether some types of factors were more strongly related to excess deaths than others. Furthermore, we could assess whether the socioeconomic signature of excess deaths changed over time during the pandemic.

## METHODS

### County-level variables

County-level variables were drawn from the raw Area Health Resources File (AHRF) (15) (in contrast to pre-built composite area-level indices such as the Area Deprivation Index (16)), downloaded from the Health Resources and Services Administration data warehouse (https://data.hrsa.gov/data/download). The released file contains 7,236 county variable columns with different columns for the same variable measured at different years. Each variable carries an 8-character feature code (f + 7 digits) whose last two digits encode the data year (e.g. ‘10 = 2010, ‘18 = 2018, ‘22 = 2022). We applied a latest-year retention rule — for each base variable appearing in several years, we focused only on the most recent eligible year in the pre-pandemic “2019–2020” file, so as to consider the most recent pre-pandemic information, leaving 3,162 assembled columns. Of these, 2,745 were AHRF predictor variables that form the input to all downstream analyses (median year of reference 2018, and only 11 variables carrying a data year of 2020, all of them administrative or geographic codes); the other 417 columns are not predictor variables — they were the mortality outcomes themselves (n=324), their population and death denominators (n=54), and geographic/administrative identifiers (n=39) (Supplementary Text; full list in excluded_columns_417.tsv). Figure S1 diagrams the full pipeline from source files to this variable set. The United States has 3,143 counties and county-equivalents in the 50 states and the District of Columbia, and the mortality database covers 3,139 of them; the four absent are three Alaska census areas and Oglala Lakota County, South Dakota, all of them products of boundary changes after 2015. Missing predictor values were filled with the mean of that variable across counties, which affects 2.15% of cells. The fill is conservative: leaving those cells missing instead raises the count of variables reaching |CC| > 0.30 from 656 to 674 (Supplementary Text; Table S1).

In two sensitivity analyses, we also considered the most recent year available for each variable (using the “2023–2024” file with median year 2022); and the most recent year up to 2015 (with median year 2015). Table S2 gives the per-data-year variable counts for the main and sensitivity analyses. Table S3 gives the same composition broken down by semantic super-cluster.

### Normalization (per-capita)

All variables were expressed per capita. Of the 2,745 predictors, 192 were already rates, percentages, or ratios (thus already per capita) and 2,526 were raw counts that were divided by the population of the respective year; the remaining 27 (county land area and a small number of administrative or metadata code variables) were not divided by population. All variables were then rescaled across counties to a 0–9,999 range, with 9,999 representing the maximum. All correlations below were computed on the normalized data.

### Mortality outcome

The outcome was the per-year excess-death proportion (asedx_p) for each year from 2020 to 2024, asedx_p = (ased / ased_bl) − 1, where ased is the county’s observed deaths in that year and ased_bl is the deaths expected from its own pre-pandemic experience. The expected count is indirectly standardized. For each single year of age from 0 to 84, and for a combined 85– and – over band, the county’s mean 2017–2019 age-specific death rate was applied to its population in that band in the index year, and the products were summed. No external standard population enters the outcome. Because the reference rates are each county’s own, differences between counties in age structure are removed by construction, as are shifts in a county’s own age structure over time. The upstream database also computes age-standardized rates against the 2000 US Standard Million, but those columns are not the outcome used here. The outcome is a proportion (e.g. 0.20 = 20% above expected). The World Health Organization declared the end of the COVID-19 public health emergency of international concern on 5 May 2023, so 2023 (in part) and 2024 are not pandemic years. We keep them because the question is how long the socioeconomic structure of excess death lasts, and the outcome returns to its pre-pandemic level by 2024 (Table S4). The outcome is all-cause excess death and not COVID-19-coded mortality, because assigning COVID-19 as the underlying cause depends on local certification and testing practice and varies between counties and over time.

A county was omitted if it recorded fewer than 10 deaths in any age group used (all ages, under 65, 65 and over) in any year from 2017 to 2024: its excess-death ratio then rests on too few deaths to be stable, and the data-use rules of the mortality source described below forbid publishing statistics based on nine or fewer deaths. This removes 361 counties holding 0.38% of the population, among them Kalawao County, Hawaii and Loving County, Texas, which recorded no deaths across the 2017–2019 baseline and whose outcome is therefore undefined. The analysis uses the remaining 2,778 counties, which hold 99.6% of the population of the United States. Two errors in the county populations that enter the expected count were corrected after submission (Supplementary Text): the 2017–2019 populations, estimated on the 2010 Census base, are now bridged to the 2020 Census base, so the denominators no longer step at 2020, and the eight Connecticut counties, which the state replaced with nine planning regions, are rebuilt from town populations.

Values were taken from a purpose-built county-level mortality database derived from the US Centers for Disease Control and Prevention WONDER underlying-cause-of-death data (17). CDC WONDER suppresses any sub-national cell holding 1 to 9 deaths; cells with no deaths were requested explicitly. At the single-year-of-age resolution the standardization requires, this withholds most of the table: of 2,162,126 county-year-age cells in 2017–2024, 1,779,631, or 82.3%, hold fewer than 10 deaths, and those cells carry 11.9% of all deaths.

These counts were recovered exactly rather than estimated. For each county the query issued to CDC WONDER was every county except that one. Such a complement aggregates more than three thousand counties, sits far above any suppression threshold, and is released in full. Subtracting it from the national total leaves the target county:

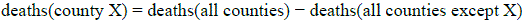

One query is run per county, so each contains exactly one unknown. Recovery is therefore exact for every county independently, and it does not degrade as the number of small counties grows. We verified that the complement is never itself suppressed: across all 9,426 complement downloads for the 3,142 counties, covering 4,486,776 data rows, no row was withheld and the smallest cell value was 14. Summing the recovered counts reproduces the published national all-cause death total in every year from 2017 to 2024 to within 0.011% (Table S5). Where a single-year age band carried a zero population estimate but a non-zero death count, the death was moved to the next band with positive population, so no death was discarded and no zero denominator was formed.

The main analysis used the all-age series; the same outcome was also computed for the under-65 (LT65) and 65– and – over (GE65) age groups, which we analyzed separately (Table S6). The suppression burden differs sharply between these: 1.0% of all-age county-years hold fewer than 10 deaths, against 7.3% of under-65 county-years.

### Association measure and effect-size bands

For each (variable, year) we computed a weighted Pearson correlation coefficient (CC) between the variable and the excess-death measure across counties, under population weighting (each county was weighted by its 2019 population, 2019 being the last pre-pandemic year, so that the weight is not itself affected by pandemic mortality or pandemic-driven migration). In sensitivity analyses, we also examined weighting by the 3/4 power and the square root of the population, as well as analyses using nonparametric Spearman correlation coefficients that do not have to assume linearity and can better accommodate extreme outliers.

We classified correlations based on their absolute value as weak (|CC| ≤ 0.30), moderate (0.30 < | CC| ≤ 0.45), and strong (|CC| > 0.45). These are descriptive categories, not tests; the permutation null described below calibrates them against chance. We preferred focusing on the size of the correlation rather than the statistical significance, because given the large sample sizes involved p-values were expected to be highly significant even for what might be very weak correlations.

To check that a screen of this size does not generate correlations of the reported magnitude by chance, we also computed an empirical null. County labels of the whole five-year excess-death block were permuted together, holding the predictors and weights at their own rows, and the screen was rerun; 10,000 permutations. Permutation was stratified by 2019 county population in 20 equal-count strata, because the excess-death ratio of a very small county rests on few deaths, and unrestricted permutation would move that counting noise onto the large counties that carry most of the weight. After the exclusions above, the spread of the ratio below 10,000 people is 1.6 times that above 10,000, and the stratified and unrestricted nulls give critical values of 0.447 and 0.420; we report the stratified null, which is the more conservative.

### Semantic clustering of variables

Variable names were embedded with a sentence-transformer neural language model (MPNet) (18, 19) and clustered using Ward’s agglomerative hierarchical clustering (Ward linkage) (20). A key step is name cleaning: before embedding we strip surface tokens that carry no subject information — age bands (_35_44), sex, patient-care/role suffixes, and Population_ / #_ / %_ / Hospital_with_ prefixes — so the embedding keyed on what is measured rather than incidental wording. For example, the six age-banded pulmonary-disease physician counts in the dataset (Pulmonary_Diseases_<35, _35_44, _45_54, _55_64, _65_74, _75+) and the eighteen psychiatry variants spanning age bands and practice roles (Psychiatry_Total, Psychiatry_Patient_Care_Office_Based, Psychiatry_Patient_Care_Hospital_Full_Time_Staff, Psychiatry_35_44, …) each collapse to a single embedded subject (“Pulmonary diseases”, “Psychiatry”); and a per-capita rate such as %_Males_40_64_without_Health_Insurance enters as “Males 40–64 without health insurance”. This process roughly doubled cluster cohesion as assessed by the silhouette coefficient — a cluster-cohesion score from −1 to 1, higher meaning tighter, better-separated clusters (21). We selected k = 120 clusters and then generated 11 super-clusters. The organizing unit for the presented analysis is the 11 super-clusters; the 120-cluster level is a finer index used for de-duplication and within-cluster ranking.

### Super-clusters

To generate the 11 super-clusters, the 120 cluster labels were themselves embedded, and a Ward linkage on the label cosine distance was cut at k = 11 to give contiguous super-clusters (renumbered top-to-bottom in dendrogram order). The 11 super-cluster names were written to match each branch’s membership.

### Reproducibility

The entire CC-dependent analysis layer regenerates from one script, code/run_standard_k120_w1.0.sh (steps A–K), writing all figures to figures_2745/ and tables to the project root and ward_sem_clean2_k120/. The k=120 clustering build, the 120 cluster labels, and the 11 super-cluster names are curated artifacts reused by the pipeline; everything downstream is scripted.

### Software

The correlation analyses, clustering, sensitivity analyses and figures were implemented in Python. Mortality-data acquisition was automated in JavaScript with Playwright and Chromium, and the mortality and population processing and validation were implemented in R. Scripts were developed iteratively with the assistance of a large language model (Claude, Anthropic) used as a coding aid: the investigators specified each analytical requirement, reviewed all generated code for correctness, and validated the outputs through multiple independent cross-checks, confirming that the |CC| moderate-association classification was stable across the 2019–2020, 2023–2024, and 2015 data vintages, and that the county death counts reproduced both the published national totals (Table S5) and, through the upstream pipeline’s own assertion, every state-year total to within 60 deaths. Variable-name embedding used the MPNet sentence-transformer; clustering used scikit-learn. The final scripts are deterministic and reproducible and are provided, with the derived result files, in the public repository (see Data and code availability).

## RESULTS

### Structure of the variable space

Cleaned-name clustering produced 120 cohesive semantic clusters (silhouette 0.42 at k=120; cluster sizes range 7–66 variables, median 18 — Figure S2). These clusters were aggregated into 11 super-clusters (Table S7; Figure 1); the full membership (each super-cluster and its constituent cluster names) is given in Table S8. No internal criterion selects k=120 or k=11: the silhouette coefficient rises monotonically from k=2 to k=1000 without a peak (Figure S3), and the largest gaps between successive dendrogram merge heights fall at k=2, 4 and 7. We chose k=120 so that clusters are small enough to serve as a de-duplication unit and large enough to carry an interpretable label, and every result is reported at the level of the 11 super-clusters.

**Figure 1.**
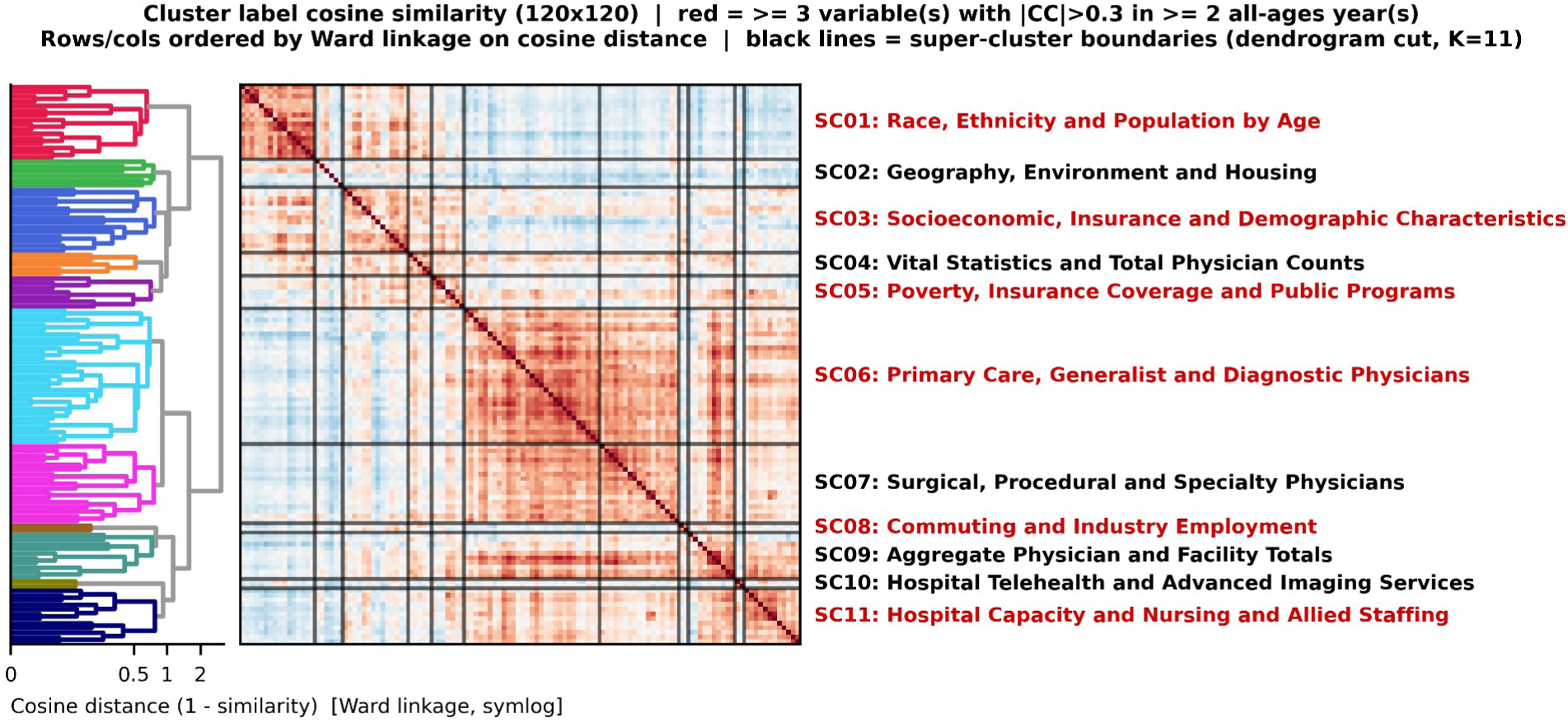
120×120 cluster-label cosine matrix with per-super-cluster-colored Ward dendrogram (symlog cosine-distance axis). Super-cluster labels are reddened when a super-cluster holds ≥3 variables each reaching the moderate band (|CC| > 0.30) in ≥2 *years.* The 120 cluster names are listed in Table S8; they are not printed on the axis because 120 labels cannot be set legibly at the printed width.

Of the 2,745 screened variables, 2,727 carried a valid CC and a semantic-cluster assignment (the basis for Table 1); the remaining 18 were constant across all counties — all-zero counts of rare physician sub-specialties or services, or administrative name fields — giving zero variance and hence non-computable correlations. Across these 2,727 variables, the distribution of the maximum |CC| for any year in the period 2020–2024 had a median of 0.181, mean 0.213, 90th percentile 0.415, 95th percentile 0.493, and maximum 0.642. Overall, 24.1% of variables (656) reached the moderate correlation level (|CC| > 0.30) in at least one year and 7.4% (202) reached the strong correlation level (|CC| > 0.45) in at least one year. These are not independent findings: the 656 variables fall into 67 of the 120 semantic clusters and 10 of the 11 super-clusters, and the 202 strongest into 29 clusters within 7 super-clusters, so the signal is a broad and redundant socioeconomic gradient rather than a set of discrete risk factors. Figure 2 shows the full per-variable CC distributions by year and age group. As shown, moderate and strong correlations (| CC| > 0.30) appeared among variables in 2020 (11.4%) and 2021 (14.8%). In 2022 only some moderate correlations were seen (3.3%) and in 2023 few (36 variables, 1.3%), while few appeared in 2024 (4.1%). Between-county dispersion of the outcome narrowed over the same years, with the population-weighted standard deviation falling from 0.103 in 2020 to about 0.055 in 2022–2024 (Table S4), but that 45% narrowing cannot produce the 4.5-fold fall in the share of variables reaching |CC| > 0.30. In the age strata, the patterns were very similar to the overall analysis, with moderate and strong correlations appearing in the first two years of the pandemic and then diminishing or even disappearing (even more prominently in the under-65 stratum) in subsequent years.

**Figure 2.**
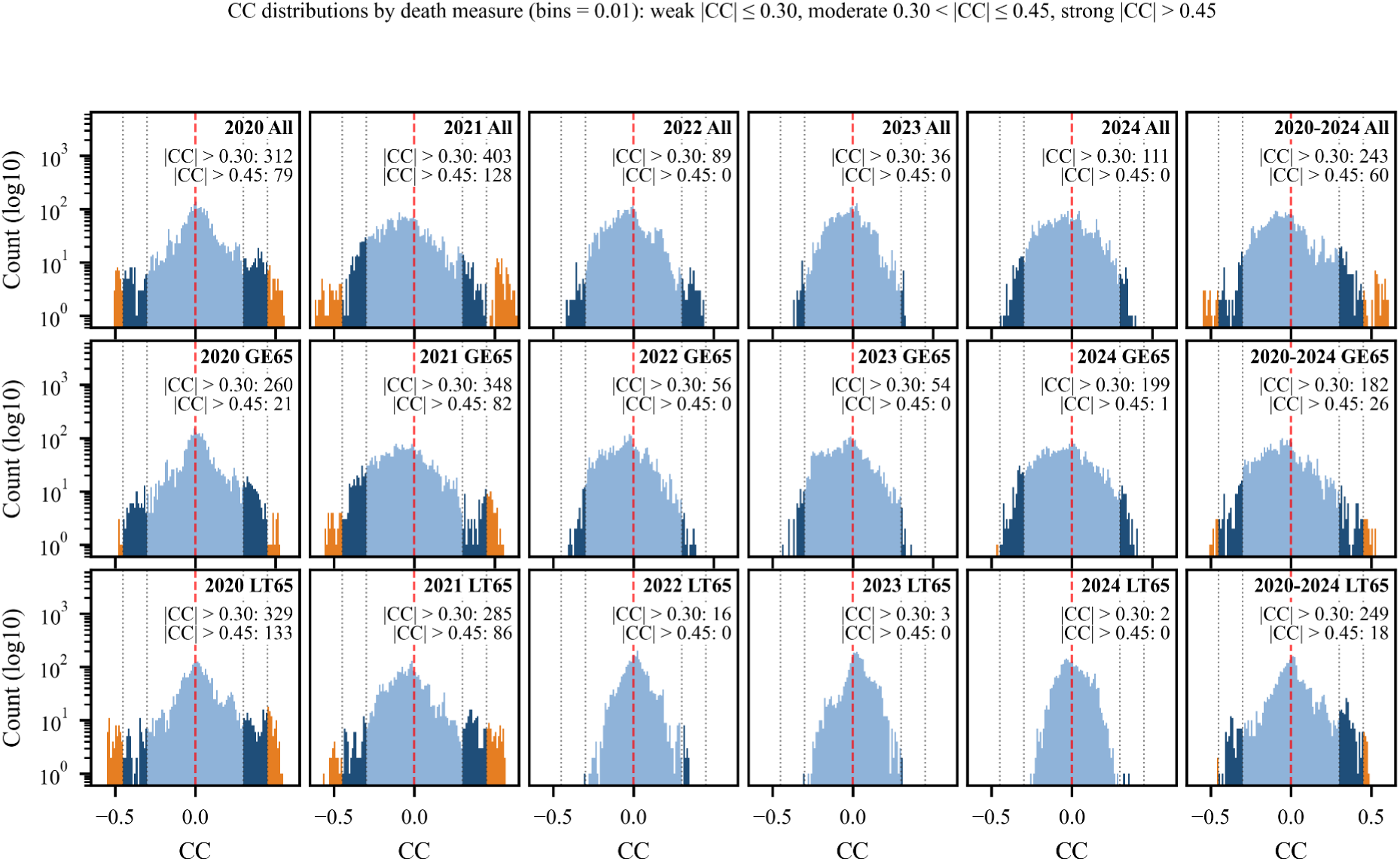
Per-variable correlation-coefficient (CC) distributions, 2019-vintage predictors, population-weighted. Columns: death years 2020–2024 plus pooled 2020–2024; rows: All / Ages ≥65 / Ages <65. log₁₀ count on the y-axis; dark blue marks moderate *(|CC| > 0.30), orange strong (|CC| > 0.45); red line at CC = 0.* Panel counts are over the 2,727 variables carrying a valid correlation, so they match the Results.

**Table 1.**
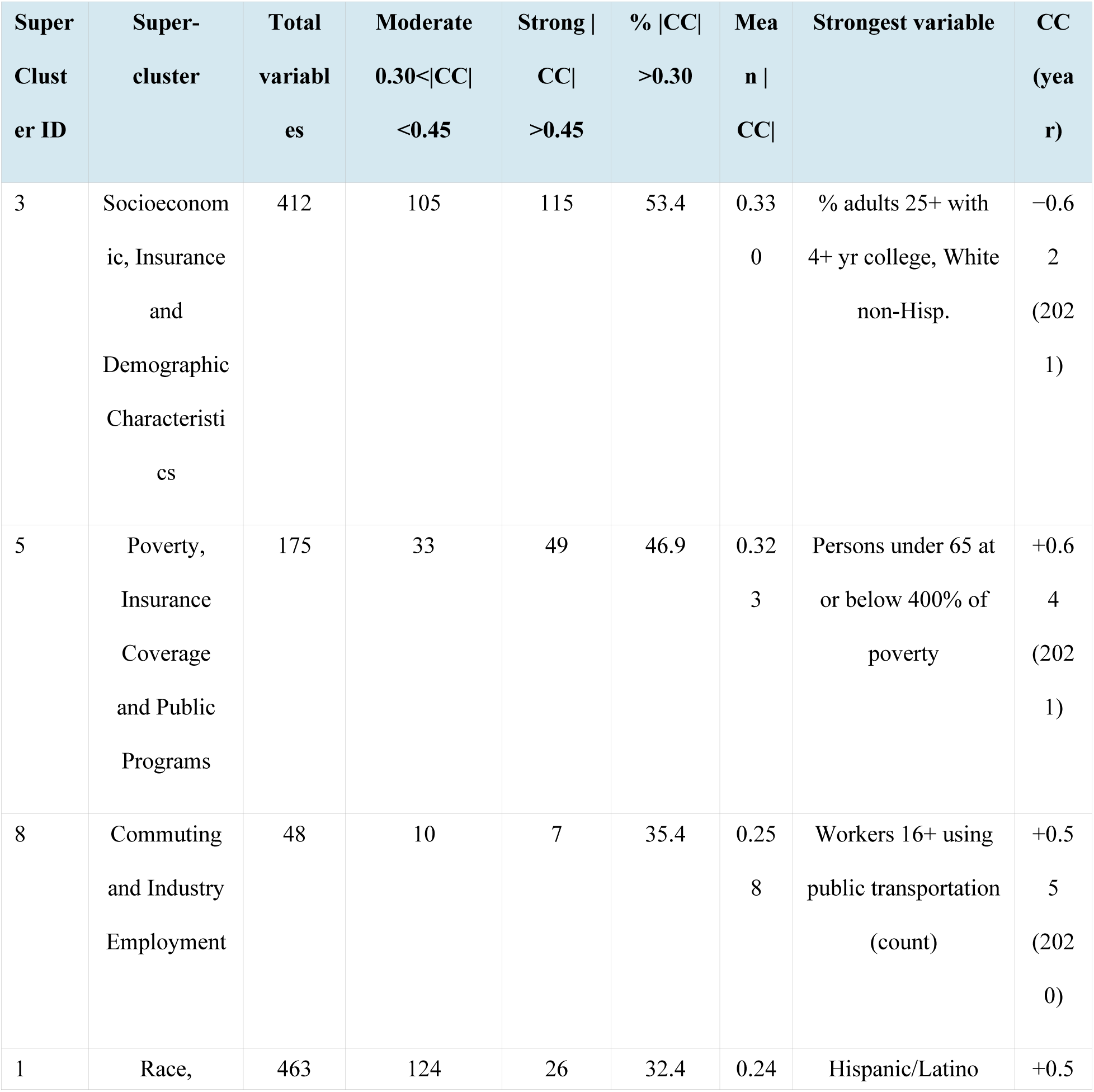
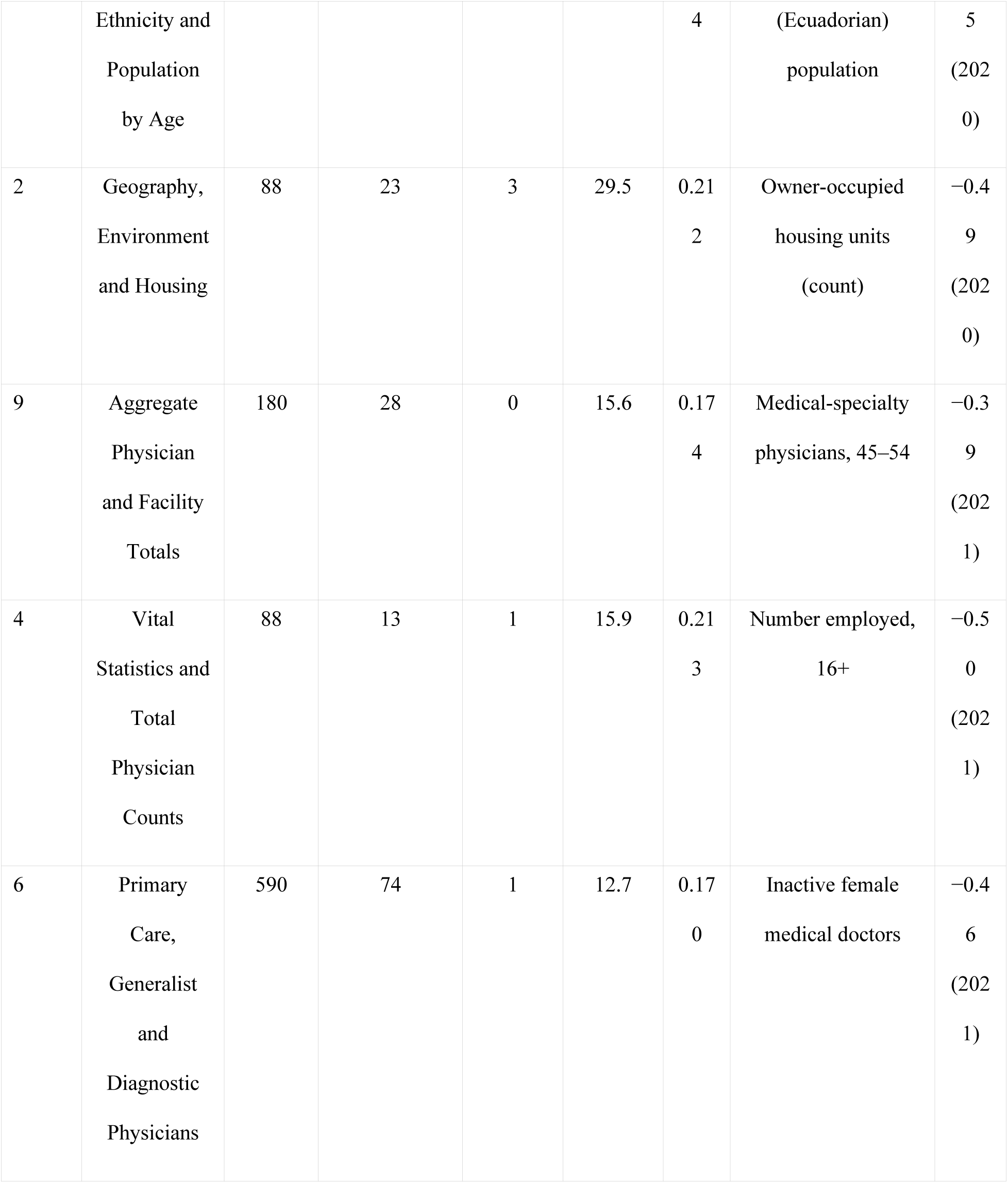
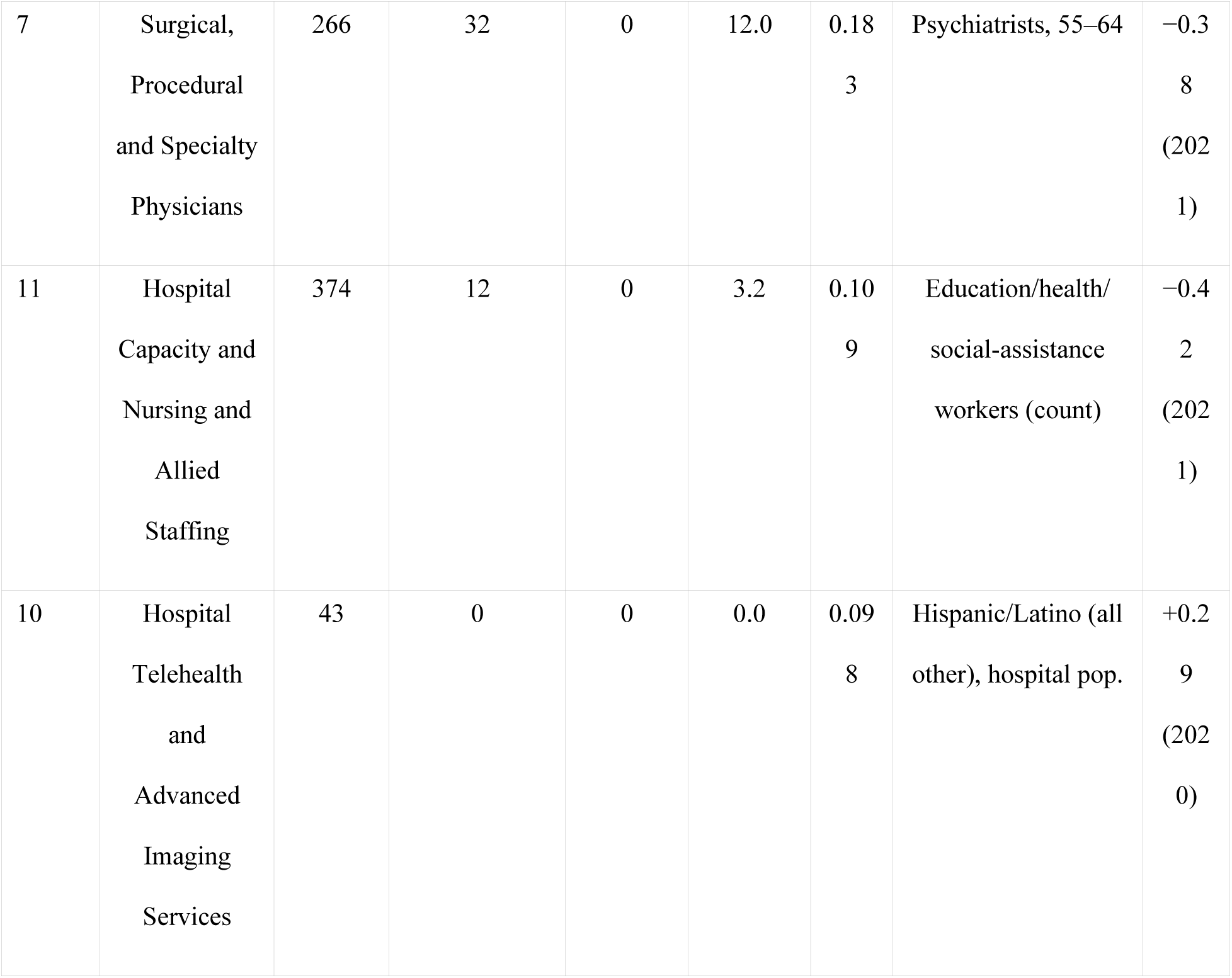
Super-cluster association summary. For each of the 11 super-clusters, the count of variables whose maximum |CC| across the all-age years 2020–2024 (population-weighted, NORMED) reaches the moderate band (0.30 < |CC| < 0.45) and the strong band (|CC| > 0.45), with the percentage attaining |CC| > 0.30 and the mean of per-variable maximum |CC| (2,727 variables with a valid CC and a semantic-cluster assignment).

The screen as a whole is far outside its empirical null (Figure S4). Under 10,000 population-stratified permutations of the excess-death outcome, a mean of 7.3 variables reach |CC| > 0.30 against the 656 observed, and no permutation reaches 656 (p = 0.0001); 44.5% of permutations produce no variable above 0.30 at all. The implied false-discovery proportion in the moderate band is about 1.1%. The 95th percentile of the null distribution of the largest |CC| anywhere in the screen is 0.447, so the 207 variables above it, including all 202 in the strong band, clear the threshold that controls the family-wise error rate at 5% across all 13,725 tests, while variables between 0.30 and 0.447 are jointly far beyond chance without being individually significant after correction.

For interpretation we group the eleven super-clusters by subject matter into two families: socioeconomic/demographic (SC1, SC2, SC3, SC5, SC8) and health system (SC4, SC6, SC7, SC9, SC10, SC11). This grouping is by content, not a data-driven partition — a two-way cut of the cluster-label dendrogram would instead split the super-clusters into SC1–5 and SC6–11 (placing the physician-count super-cluster SC4 with the demographic block and employment SC8 with the workforce block). Our conclusion does not depend on that judgement: under the purely algorithmic two-way cut, 40.1% of the 1,226 variables on the socioeconomic side reach | CC| > 0.30 against 10.9% of the 1,501 on the health-system side, against 41.7% and 10.4% for the content-based grouping used here.

### Socioeconomic and demographic factors

The variables with moderate or strong correlation were predominantly found in the demographic and socioeconomic family (Table 1). These five super-clusters — SC1 (Race, Ethnicity and Population by Age), SC2 (Geography, Environment and Housing), SC3 (Socioeconomic, Insurance and Demographic Characteristics), SC5 (Poverty, Insurance Coverage and Public Programs) and SC8 (Commuting and Industry Employment) — comprise 1,186 variables, 42% of which reach |CC| > 0.30; they account for 295 of the 454 variables in the moderate band and 200 of the 202 variables reaching the strong band.

The strongest correlations attained were with poverty share (CC +0.64 in 2021), college attainment (−0.62 in 2021), and high income (−0.61 in 2021) in the overall analysis. In the ≥65 stratum the three strongest were two college-attainment measures and a poverty-share measure (−0.55, +0.55 and −0.55, all in 2021); in the <65 stratum they were poverty share (+0.57), uninsurance among women aged 40–64 (+0.56), and high household income (−0.56). The top six distinct moderately associated variables in each super-cluster (deduplicated to one per semantic cluster) are listed in Table S9, and the highest-|CC| variable in every one of the 120 clusters is given in the full per-cluster table (top_metric_per_cluster_2020_2024_w1.0.tsv, provided as Supplementary Data).

To show where the affected counties are, we averaged the 200 strong-band socioeconomic and demographic variables into a single advantage index, each standardized across counties and sign-aligned so that a high score means lower excess death. The index adds no analyst choice, because its members are the ones the screen selected. In population-weighted quintiles of about 65 million people each, excess death in 2020 and 2021 falls monotonically from 0.274 in the most disadvantaged fifth to 0.136 in the most advantaged, and the gradient persists into 2022 to 2024 at a fifth of that level (Table S10). Geographically the extremes are the West South Central division at 0.241 and New England at 0.136 (Table S11; Figure 3). Rurality is not the explanation: the rural population share is 13.0% in the most disadvantaged fifth and 20% to 23% in each of the other four.

**Figure 3.**
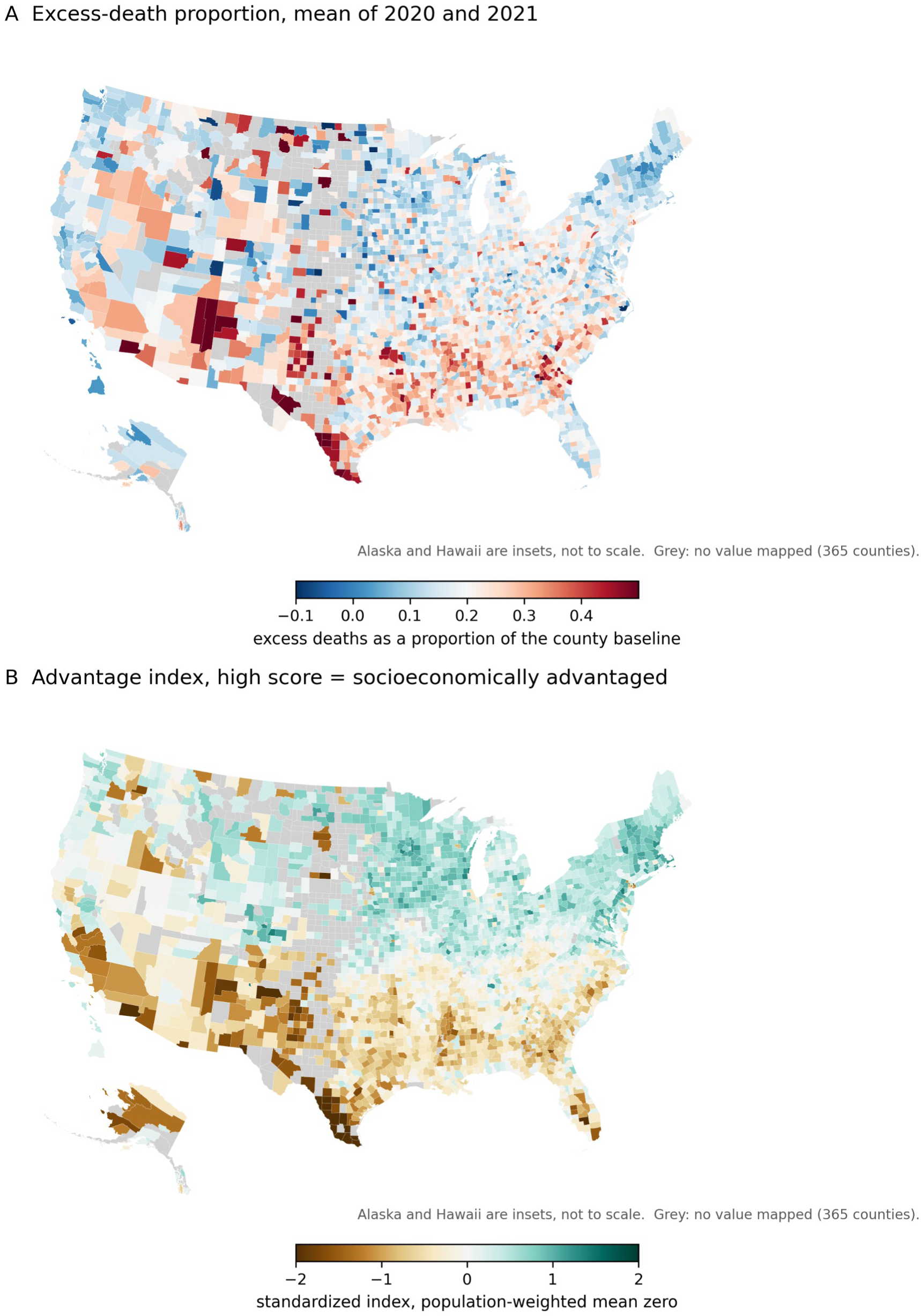
County maps of the outcome and of the advantage index, 3,143 counties in an Albers equal-area projection with Alaska and Hawaii shown as insets and not to scale. (A) Excess-death proportion, the mean of 2020 and 2021, on a scale centred *at 0.20. (B) The advantage index of Table S10, the mean of the 200 socioeconomic and demographic variables reaching |CC| > 0.45, standardized and sign-aligned so that a high score corresponds to lower excess death. The two panels show the same geography: the band of high excess death through Texas, the lower Mississippi valley and the southeast coincides with the band of low advantage, and New England and the upper Midwest are light in panel A and dark in panel B. Grey counties carry no value: four because the mortality database does not cover them and 361 because they are omitted from the analysis (Methods)*.

Super-cluster 4 holds the pre-pandemic cause-specific and age-specific death measures, among them infant deaths, deaths from suicide, homicide and motor vehicle accidents, and mortality counts by age band. The strongest of them reaches only |CC| = 0.24, but this is not evidence that prior mortality is unrelated to pandemic excess death: AHRF withholds each of these counts below 10, so for the rarer events almost every county is withheld, and the variables carry little information between counties.

The pattern was internally consistent: counties with higher affluence, education and insurance coverage had lower excess death, and counties with more poverty, uninsurance, disability, non-employment and larger shares of certain minority populations had higher excess death. Given that no single variable exceeded |CC| ≈ 0.64, the largest squared correlation is 0.64² = 0.41, so under 42% of the cross-county variation in excess death is shared with any single variable (Figure S5).

### Health system factors

Across the 1,541 health system variables — the majority of the catalog (super-clusters 4, 6, 7, 9, 10, 11: physician counts, the physician workforce, and hospital capacity and facilities) — only 10.4% reached |CC| > 0.30 (with hospital-capacity and telehealth/imaging variables at 0–3%) versus 42% of the 1,186 socioeconomic and demographic variables (super-clusters 1, 2, 3, 5, 8) (Table 1). Only two health system variables reached a strong correlation: the number employed aged 16 and over, which the clustering placed in super-cluster 4 (−0.50), and inactive female doctors (−0.46). The six health system super-clusters contributed the bulk of the catalog (75 of 120 clusters) but little of the top signal. Physician-density variables correlated negatively with mortality but most of these correlations were weak. This is consistent with health system workforce supply tracking the same affluence/urbanicity gradient that drives the demographic signal, rather than an independent protective association.

### Robustness in sensitivity analyses using different variable years

Variables were matched across vintages by base code (the f-code minus its last two-year digits): 2,392 base-matched between the main analysis and the older-data sensitivity analysis (latest year ≤ 2015) and 2,750 between the main analysis and the more-recent-data sensitivity analysis (the AHRF 2023–2024 release). Many matched variables were, however, identical by construction — their in-use vintage already qualified, so the backward cutoff or the forward value-swap left them unchanged — and such variables would be uninformative for a robustness test; we therefore restricted the comparison to the variables whose vintage value actually changed (1,617 for the ≤2015 comparison and 1,971 for the 2023–2024 comparison; the 775 and 779 identical variables are excluded). Among these genuinely refreshed variables the per-variable correlation with excess death was highly stable: each variable’s CC (2020–2024) in the main analysis tracked the older-data value at Spearman ρ = 0.97 (Pearson 0.97) and the more-recent-data value at ρ = 0.95 (Pearson 0.96); the moderate-association (|CC| > 0.30) classification agreed 97.5% and 97.0% (Table S12); and among the 397 and 399 variables reaching |CC| > 0.30 in the main analysis, three reversed sign with the older data and two did with the more recent data (Figure S6). Full distributions of correlations appear in Figure S7 and Figure S8.

### Sensitivity analysis on population weighting

To assess sensitivity to the weighting choice, we recomputed all correlations across a range of weight powers (each county weighted by its 2019 population raised to the power p; Table S13). Poverty-share, uninsurance and college-attainment measures led at every weighting, although their order varied, and the magnitudes scaled smoothly with p. At p = 0.75, a middle ground that roughly halves the dominance of the largest counties (Kish effective sample size (22) 695, versus 279 at full weighting), 18.1% of variables reached a max |CC| > 0.30 and 4.4% the strong band, versus 24.1% and 7.4% under the primary full-population weighting; the single strongest correlation was 0.59 versus 0.64. Square-root weighting (p = 0.5) brought the signal down to 11.8% moderate and 2.6% strong, against 5.6% and none unweighted. The headline effect sizes are therefore sensitive to how strongly large metropolitan counties are weighted, although the qualitative pattern — socioeconomic and demographic variables dominating, health system variables being weaker — holds at every weighting. Table S14 sets out which aspects of these results are robust and which are not.

### Sensitivity analysis on the correlation method

The semantic super-cluster structure was unaffected by use of Spearman instead of Pearson correlation coefficients. The leading correlates were again poverty-share and uninsurance measures. Spearman correlations were larger at the top and in the moderate band but not in the strong band: the single strongest correlation rose from |CC| = 0.64 to 0.67, the share of variables reaching a max |CC| > 0.30 rose from 24.1% to 32.3%, and strong correlations fell from 7.4% to 6.2% (Table S15). The qualitative pattern was preserved: socioeconomic and demographic variables again dominated, and the health system super-clusters remained far weaker — two variables reached the strong correlation level under Pearson (one each in super-clusters 4 and 6), and three under Spearman (one each in super-clusters 4, 6 and 11). Table S16 lists the twenty strongest correlations with both coefficients side by side.

## DISCUSSION

We have evaluated at large scale the correlational footprint of 2,745 county-level variables against excess deaths during 2020–2024. Besides demographics, the social-determinant gradient that predicts many adverse health outcomes (with education (23), income (24), and insurance (25) being protective and poverty, uninsurance, disability, non-employment, and specific minority-population shares being risk-associated) offered the strongest signals for associations with excess deaths during 2020 to 2024. Health system supply and related variables (26) added a weak, mostly protective signal. Overall, even the strongest single variable topped out at |CC| ≈ 0.64; therefore, no county characteristic is close to deterministic, and the most honest summary may be the full-scale presentation of a multifactor socioeconomic gradient rather than any one specific factor.

A standing critique of ecological COVID studies is that associations are fragile to data choices. This problem largely jeopardizes studies that claim to identify highly specific predictors as being more important. However, in our analysis, the big picture where thousands of variables are considered remained robust to different analytical choices. Moreover, we demonstrated that results did not change substantially when we considered earlier or later measurements for the variables. This may reflect the fact that most of the county-level features do not change substantially over time. Disadvantaged counties remain disadvantaged and affluent ones remain affluent, with relatively few exceptions. Socioeconomic factors remain pervasively associated with health and disease outcomes. The socioeconomic signal also faded as the pandemic moved past its first two years. This is not a consequence of excess death itself shrinking: a correlation coefficient is scale-invariant, so a proportional reduction in the outcome leaves it unchanged. The level of excess death did fall over the same period, reaching approximately zero in aggregate by 2023–2024 (27), and the socioeconomic structure of that excess weakened. This socioeconomic disadvantage was most important in the early phase of the pandemic before vaccines were available and before the emergence of less lethal Omicron variants. The weaker correlations seen with health system factors may suggest that socioeconomic disadvantage may be more important than the exact capacity, resources, and specialization of the health system. People who are uninsured and lack access and those who suffer from marginalization may not be able to benefit much from improvements in the health system at the hospital level and workforce (28). Socioeconomic factors operate at a deeper root than can be fixed by traditional specialist hospital care and require special solutions and serious commitment that is usually lacking (29). We should acknowledge, however, that the health system variables that we used reflected the baseline structural endowment rather than real-time surge or ICU strain.

These patterns have practical uses that need no causal claim. A county position on the socioeconomic gradient predicted its excess death in 2020 and 2021 far better than any measure of health-system capacity, so preparedness planning that ranks counties by hospital beds or physician supply will not rank them by risk. The strongest single variable reached |CC| of about 0.64, so no one characteristic identifies risk on its own and a composite of many correlated measures is the honest instrument. The variables carrying the signal here are also the ones worth collecting at individual level in studies that can address causation.

There is increasing interest in assessing the external exposome, the totality of exposures and potential risk factors to which people are exposed beyond their own genetics (30–33). Most applications examine traditional environmental exposures but may enrich the analysis with other determinants as well. These studies may vary in their selection of variables, use of individual versus group-level data or both, causality aspirations or lack thereof, and targeted outcomes. Regardless, they may offer some useful insights that extend beyond what analyses of single or few select determinants may provide. To our knowledge no previous study has examined such a large number of county-level variables in relationship to overall excess mortality. Given their complexity but also bird’s-eye view, ecological external exposome studies of this genre may prove particularly useful in offering high-level pictures about the potential architecture of risk of major outcomes (34). They may then be followed by targeted, more specific studies on factors or clusters of factors of particular interest and triangulation of evidence with additional methods (35,36). The clustering approach that we used offered some advantages in dealing with the very wide space of almost 3,000 variables. The super-clusters further allowed a more facile naming of the organized patterns of variables and high-level comparisons of different types of factors. However, it does have some limitations. A few variables may have been imperfectly placed, and the k=120 and k=11 choices were analyst decisions.

Several limitations should be discussed. The main limitation of this analysis is its ecological nature. Due to potential ecological fallacy (37), individual-level inferences should be avoided and causation claims with specific factors would be precarious. The patterns we report are therefore hypotheses generated by ecological data rather than evidence about individuals. We argue, nevertheless, that once this is fully acknowledged, obtaining such a bird’s-eye view picture may be preferable to trying, in vain, to de-confound thousands of highly correlated and confounded factors. Socioeconomic risk factors travel together and effective interventions to fix them may affect many of them concurrently. We should acknowledge that reverse causality is also possible, especially in the sensitivity analysis that used county-level variables obtained during and after the pandemic. Moreover, we did not use statistical significance testing as this would have yielded a huge number of nominally significant results of questionable meaning. The magnitude of the correlations is more meaningful. Their exact magnitude nevertheless depends on the nature of population weighting; and on whether linearity is assumed or not. Spearman correlations tended to be modestly stronger than Pearson correlations, which may reflect non-linear associations for some of the variables. We should also acknowledge that our approach would not necessarily detect associations that are U-shaped, inverse U-shaped or best-fit to other non-monotonic patterns (38). It is possible that some variables, especially health system-related ones, may exhibit such non-monotonic relationships. For example, poor individuals may suffer because of underuse of health care (39), while very wealthy individuals may overuse resources and thus have a risk of iatrogenic harms (40) and even addiction (41). However, for the vast number of informative relationships, monotonic assumptions are probably reasonable.

Acknowledging these caveats, we suggest that the study of socioeconomic and health system determinants of health would benefit from additional large-scale, bird’s-eye view analyses of comprehensive sets of many variables in diverse populations and settings. Comparison of the emerging patterns across different countries, time periods, and settings and with different robustness checks may highlight which factors are the most robust and persistent in relation to major outcomes of interest.

## Data and code availability

[https://github.com/michaellevitt/SCE-county-metrics] The county mortality series was produced with the Mortality.Watch county pipeline at commit 2788f21be8492786cd3e34608b98fa379728dbc7 (29 September 2026). The recovered county death counts, and rates computed from them, are not redistributed, in keeping with the CDC WONDER data-use restrictions; the pipeline regenerates them from CDC WONDER.

## Data Availability

All data and code used in this study are publicly available. The analysis code and processed datasets are available at https://github.com/michaellevitt/SCE-county-metrics. Source mortality data were derived from the CDC WONDER Underlying Cause of Death database, and county-level predictor variables were obtained from the Area Health Resources File (AHRF).

https://wonder.cdc.gov/deaths-by-underlying-cause.html

https://data.hrsa.gov/topics/health-workforce/ahrf

https://data.hrsa.gov/data/download

## Acknowledgments

None

## SUPPLEMENT

### Supplementary Text

Significance mode. A p-value / log-probability (LP) mode exists in the pipeline but is dormant; the project default throughout this paper is the fixed |CC| effect-size rule (moderate |CC| > 0.30; strong |CC| > 0.45), a magnitude threshold rather than a statistical-significance test.

Excluded columns. Assembly retains 3,162 columns, of which 2,745 are AHRF predictor variables with a plain-English description — the input to all analysis. The remaining 417 columns are not predictor variables and were never candidates for the semantic or correlation analysis: 324 are the mortality / excess-death outcome series themselves (asedx_p and the analogous ASMR, CMR and life-expectancy measures, across years and age bands), 54 are the population and death denominators used to construct those outcomes, and 39 are geographic / administrative identifiers (FIPS and entity codes, CBSA and region codes, land area, and urban/rural population, housing and block tallies). No AHRF predictor variable was dropped for lacking a description. The full list of all 417 is provided as Supplementary Data (excluded_columns_417.tsv).

Suppression and recovery. CDC WONDER withholds any sub-national cell of 1 to 9 deaths; cells with no deaths were requested explicitly. Because the outcome is standardized over single years of age, the affected fraction is large: 1,779,631 of 2,162,126 county-year-age cells in 2017 –2024, or 82.3%, carrying 11.9% of all deaths. By age group, the share of cells below 10 deaths is 1.0% for the all-age series, 1.4% for ages 65 and over, and 7.3% for ages under 65. Every one of these cells is recovered exactly by the complement-query difference described in Methods, so the reconstruction introduces no estimation error. As a sensitivity analysis we removed the 50 counties whose all-age annual total is itself below 10, which are the counties where the recovery does most work and where the excess-death ratio is least stable. In the analysis as first submitted, variables reaching |CC| > 0.30 rose from 472 to 526 and those reaching |CC| > 0.45 from 77 to 121, almost all of it from two of the 50 counties, Kalawao County and Loving County. The minimum-count rule described in Methods now removes all 50, together with every other county where the recovery does most work.

Two counties with an undefined outcome. Kalawao County, Hawaii and Loving County, Texas recorded no deaths across the 2017–2019 baseline, so the expected count is zero and the excess-death proportion is undefined. In the analysis as first submitted the correlation code dropped them in 2022 to 2024, where the value is infinite, but in 2020 and 2021 the source value was missing rather than infinite, so the mean fill supplied 0.18401 and 0.22876 and both counties were retained. Their combined weight was 7.6 parts in 10 million, yet excluding both from every year raised the count reaching |CC| > 0.30 from 472 to 520 and the count reaching |CC| > 0.45 from 77 to 117, through filled-in predictor values in Kalawao described in the next paragraph. The minimum-count rule now removes both.

Filling of missing predictor values. Missing numeric cells were filled with the mean of the variable across counties, which affects 164,156 of 7,625,610 predictor cells, or 2.15%, in 412 of the 2,745 variables. For 59 variables the blanks are AHRF small-count suppression, which withholds any vital-statistics count below 10, so the fill assigns a high value where the true value lies between 0 and 9. Those variables are uninformative by construction: the filled share reaches 97.8% of counties for deaths at ages 1 to 4 and 96.9% for three-year Black male infant deaths. Of the 656 variables reaching |CC| > 0.30, three have more than 5% of their cells filled, the largest 5.40%, and none of the 202 reaching |CC| > 0.45 has more than 0.86%. In the analysis as first submitted the fill mattered almost entirely through one county. Kalawao County, Hawaii has 85 residents. Where its value for a count variable was blank, the fill supplied the national mean county count, typically thousands to tens of thousands, and the per-capita step divided it by 85. For persons under 65 at or below 400% of poverty the fill supplied 51,499, which becomes 606 such persons per resident, against 0.85 in the largest real county. In 92 variables a filled-in Kalawao value of this kind supplied more than half the weighted variance, and in 46 of them more than 90%, pulling the correlation toward zero: for the poverty measure above, from 0.561 to 0.168 in 2021. The minimum-count rule removes Kalawao. In the reported analysis, leaving every filled-in cell missing, and setting aside the 83 variables whose real data cover less than 90% of the population, raises the count reaching |CC| > 0.30 from 656 to 674 and the count reaching |CC| > 0.45 from 202 to 211 (Table S1). Every treatment that keeps the full variable set gives at least as many variables above both thresholds as we report, so the reported analysis is the conservative one.

Corrections to the mortality extract after submission. The county series was regenerated in September 2026 with four corrections, and every result in this paper was recomputed on it. First, Connecticut replaced its eight counties with nine planning regions, and from 2020 the population estimates in the submitted extract followed the regions while the deaths still followed the old counties: Middlesex County fell from 163,790 people in 2019 to 65,321 in 2020, which put its excess death at +162% in 2020. The eight counties are now rebuilt from the planning regions with the Census town-to-region correspondence and April 2020 town populations; Middlesex has 163,578 people in 2020 and an excess of +13.6%. Second, county populations for 2017 to 2019, estimated on the 2010 Census base, had been joined to estimates on the 2020 Census base with only a state-level adjustment, so the expected count of a county could step at 2020. They are now bridged county by county, with each county total the sum of its single-year-of-age populations, and outside Connecticut the number of counties whose population changes by more than 5% between 2019 and 2020 falls from 475 to 55. Third, final 2024 death data replace the provisional data of February 2026, which closes the 2024 gap in Table S5. Fourth, the all-race series is built from all-race queries rather than summed from race-specific cells. Outside Connecticut these corrections change the excess-death proportion of an analyzed county-year by a median of 1.6 percentage points (95th percentile 7.5). The eight Connecticut counties, whose outcome was wrong in the submitted analysis, are now correct and are included.

### Supplementary Data

Seven machine-readable files accompany the manuscript:

1. **Top_metric_per_cluster_2020_2024_w1.0.tsv** (the highest-|CC| variable in every one of the 120 clusters, 2020–2024);
2. **Table_S2_vintage_composition.tsv** (variable counts by data-year for each AHRF release, rendered as Table S2);
3. **Table_S6_age_band_divergence.tsv** (moderate-association counts and max |CC| by year and age band, rendered as Table S6);
4. **Table_S8_supercluster_cluster_names.tsv** (the full super-cluster → cluster membership listing, also rendered as Table S8 below);
5. **Table_S9_top6_metrics_per_supercluster.tsv** (the top six distinct moderately associated variables per super-cluster, rendered as Table S9);
6. **Table_S9A_collapsed_variants.tsv** (every near-duplicate variable removed in the Table S9 deduplication, listed under its representative); and
7. **Excluded_columns_417.tsv** (the 417 non-predictor columns excluded from analysis).

### Supplementary Tables

**Table S1.**
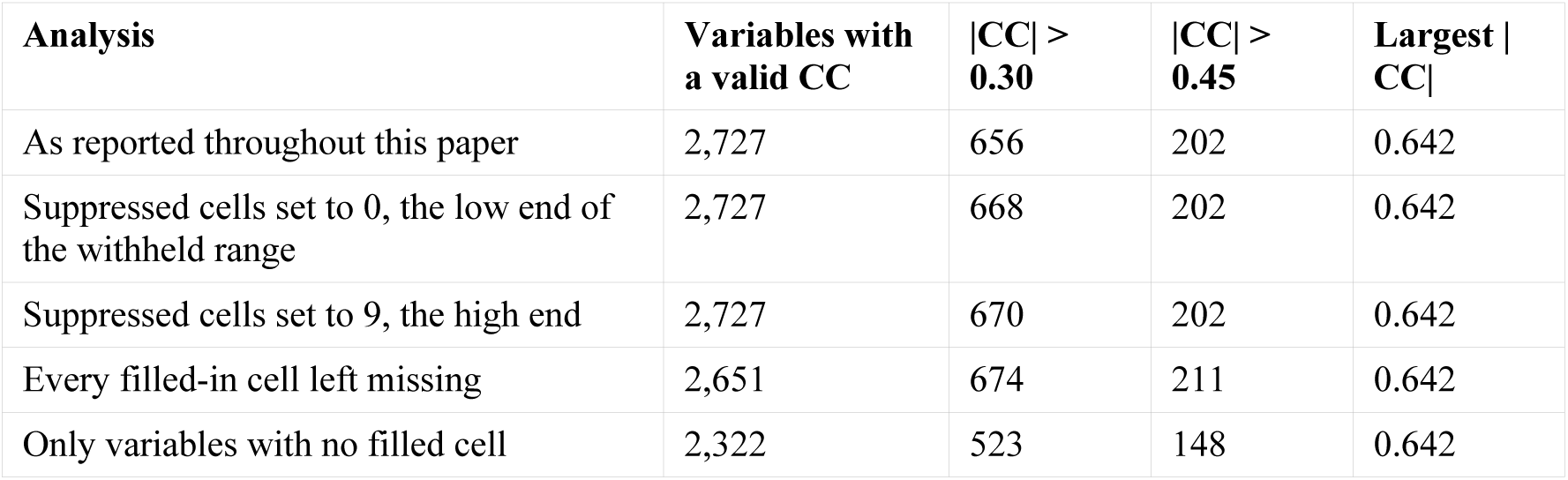
Sensitivity of the screen to the treatment of missing predictor values described in Supplementary Text. The first row is the analysis reported throughout the paper. The suppressed cells are the AHRF vital-statistics counts below 10, withheld at source and filled with the column mean; setting them to 0 and to 9 brackets their true range. When every filled-in cell is left missing, variables whose real data cover less than 90% of the population are set aside, since otherwise a variable observed in a handful of counties is correlated on a handful of points; this removes 83 variables. The third and fourth columns are counts of variables, not percentages. Every alternative that keeps the full variable set gives at least as many variables above both thresholds as the analysis we report, so the reported figures are the conservative ones.

| Analysis | Variables with a valid CC | \|CC\| > 0.30 | \|CC\| > 0.45 | Largest \|CC\| |
| --- | --- | --- | --- | --- |
| As reported throughout this paper | 2,727 | 656 | 202 | 0.642 |
| Suppressed cells set to 0, the low end of the withheld range | 2,727 | 668 | 202 | 0.642 |
| Suppressed cells set to 9, the high end | 2,727 | 670 | 202 | 0.642 |
| Every filled-in cell left missing | 2,651 | 674 | 211 | 0.642 |
| Only variables with no filled cell | 2,322 | 523 | 148 | 0.642 |

**Table S2.** Data-year composition of the variables actually used in each analysis, after the latest-year retention rule (one most-recent year retained per variable). Columns: the ≤2015 backward-cutoff vintage, the 2019–2020 main analysis, and the 2023–2024 forward vintage; each row gives the number of retained variables carrying that data year (f-coded predictors plus the non-f-code count, rate and land-area predictors; pure geographic/administrative identifier codes are excluded). Variables carrying no data year at all, such as contiguous-county identifiers and census region and division codes, were previously pooled into the 2020 row; they are now shown separately as “No data year”, and the 2020 row holds only variables genuinely dated 2020. The 2019–2020 column is dominated by 2018, and secondarily 2010 and 2014 years; the 2023–2024 column is dominated by year 2022. Totals 2,395 / 2,754 / 2,754. The 2019–2020 file includes only ≤2019 variables, except for 40 variables from 2020: 29 are 2020 decennial-Census predictors — county population, housing-unit and land-area counts together with their densities and urban/rural shares, for which the 2020 Census is the most recent available measurement — and 11 are administrative geographic-identifier codes and names (Core-Based and Combined Statistical Area and Metropolitan Division labels), several of the latter being among the zero-variance variables dropped from the analysis.

| Data year | $\leq 2015$ vintage | 2019–2020 vintage<br>(main analysis) | 2023–2024 vintage |
| --- | --- | --- | --- |
| 1960 | 1 | 1 | 1 |
| 1970 | 1 | 1 | 1 |
| 1976 | 1 | 1 | 1 |
| 1990 | 1 | 1 | 1 |
| 2000 | 7 | 7 | 7 |
| 2001 | 2 | 2 | 2 |
| 2003 | 13 | 13 | 13 |
| 2004 | 4 | 4 | 4 |
| 2010 | 589 | 589 | 589 |
| 2012 | 11 | 11 | 11 |
| 2013 | 19 | 19 | 18 |
| 2014 | 345 | 345 | 345 |
| 2015 | 1361 | 12 | 12 |
| 2016 | 0 | 71 | 68 |
| 2017 | 0 | 3 | 1 |
| 2018 | 0 | 1539 | 6 |
| 2019 | 0 | 95 | 2 |
| 2020 | 11 | 11 | 3 |
| 2021 | 0 | 0 | 78 |
| 2022 | 0 | 0 | 1456 |
| 2023 | 0 | 0 | 87 |
| 2024 | 0 | 0 | 19 |
| No data year | 29 | 29 | 29 |
| Total | 2395 | 2754 | 2754 |

**Table S3.** Data year of the predictor variables by semantic super-cluster, main analysis. Counts are the full 2,745 predictors, so they exceed the Table S7 counts by the 18 variables with no usable correlation. The data year is taken from the variable’s feature code and agrees with the year in its description for all 2,725 variables that carry one. The demographic and socioeconomic super-clusters that supply the strongest correlations are the older measurements, largely 2010 census and 2014 American Community Survey data, while the weakly correlated health-system super-clusters are the more recent. The stronger associations therefore rest on information measured further before the pandemic.

| Super-cluster | Variables | Median data year | Range | % dated 2019 or later |
| --- | --- | --- | --- | --- |
| 1 Race, Ethnicity and Population by Age | 463 | 2010 | 2010–2018 | 0.0 |
| 2 Geography, Environment and Housing | 94 | 2013 | 1960–2020 | 14.9 |
| 3 Socioeconomic, Insurance and Demographic | 412 | 2014 | 2010–2019 | 1.0 |
| 4 Vital Statistics and Total Physician Counts | 88 | 2016 | 2010–2019 | 5.7 |
| 5 Poverty, Insurance Coverage and Public Programs | 176 | 2018 | 2010–2019 | 15.9 |
| 6 Primary Care, Generalist and Diagnostic Physicians | 597 | 2018 | 2016–2019 | 0.3 |
| 7 Surgical, Procedural and Specialty Physicians | 270 | 2018 | 2018 only | 0.0 |
| 8 Commuting and Industry Employment | 48 | 2014 | 2014–2018 | 0.0 |
| 9 Aggregate Physician and Facility Totals | 180 | 2018 | 2018 only | 0.0 |
| 10 Hospital Telehealth and Advanced Imaging Services | 43 | 2018 | 2010–2018 | 0.0 |
| 11 Hospital Capacity and Nursing and Allied Staffing | 374 | 2018 | 2001–2020 | 14.2 |

**Table S4.**
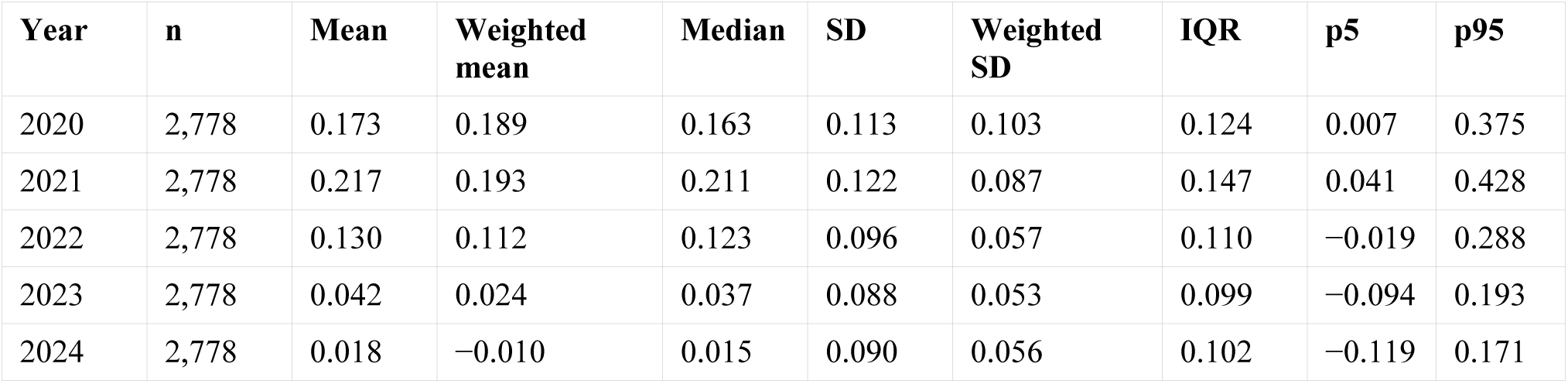
Distribution of the age-standardized excess-death proportion by year. Weighted statistics use the 2019 county population, as in the main analysis. SD is the standard deviation and IQR the interquartile range. Between-county dispersion narrows after 2021, with the population-weighted standard deviation falling from 0.103 to about 0.055, a reduction of roughly 45%. Because a correlation coefficient is scale-invariant, this narrowing does not by itself attenuate the correlations, and it is far too small to account for the 4.5-fold fall in the share of variables reaching |CC| > 0.30.

| Year | n | Mean | Weighted mean | Median | SD | Weighted SD | IQR | p5 | p95 |
| --- | --- | --- | --- | --- | --- | --- | --- | --- | --- |
| 2020 | 2,778 | 0.173 | 0.189 | 0.163 | 0.113 | 0.103 | 0.124 | 0.007 | 0.375 |
| 2021 | 2,778 | 0.217 | 0.193 | 0.211 | 0.122 | 0.087 | 0.147 | 0.041 | 0.428 |
| 2022 | 2,778 | 0.130 | 0.112 | 0.123 | 0.096 | 0.057 | 0.110 | -0.019 | 0.288 |
| 2023 | 2,778 | 0.042 | 0.024 | 0.037 | 0.088 | 0.053 | 0.099 | -0.094 | 0.193 |
| 2024 | 2,778 | 0.018 | -0.010 | 0.015 | 0.090 | 0.056 | 0.102 | -0.119 | 0.171 |

**Table S5.** Validation of the reconstructed county death counts against published national totals. County sums are the total over all 3,139 counties of the recovered all-age death counts. Published totals are the resident-death totals reported in the corresponding NCHS Data Brief, identified in the final column. The shortfall is stable near 300 deaths a year and does not grow with the total; a reconstruction that dropped or mis-assigned suppressed cells would produce an error that scaled with the 1.78 million recovered cells and varied between years. These are the deaths recorded under three historical county codes, Valdez–Cordova and Wade Hampton in Alaska and Shannon County in South Dakota, which cannot be joined to the current county geography; in 2024 they account for all 290. Sources: NCHS Data Briefs 328, 355, 395, 427, 456, 492, 521 and 548, “Mortality in the United States” for data years 2017 through 2024 respectively (National Center for Health Statistics, Hyattsville, MD).

| Year | County sum | Published national total | Difference | Percent | NCHS Data Brief |
| --- | --- | --- | --- | --- | --- |
| 2017 | 2,813,224 | 2,813,503 | −279 | −0.010% | 328 |
| 2018 | 2,838,932 | 2,839,205 | −273 | −0.010% | 355 |
| 2019 | 2,854,570 | 2,854,838 | −268 | −0.009% | 395 |
| 2020 | 3,383,392 | 3,383,729 | −337 | −0.010% | 427 |
| 2021 | 3,463,867 | 3,464,231 | −364 | −0.011% | 456 |
| 2022 | 3,279,521 | 3,279,857 | −336 | −0.010% | 492 |
| 2023 | 3,090,665 | 3,090,964 | −299 | −0.010% | 521 |
| 2024 | 3,072,376 | 3,072,666 | −290 | −0.009% | 548 |

**Table S6.**
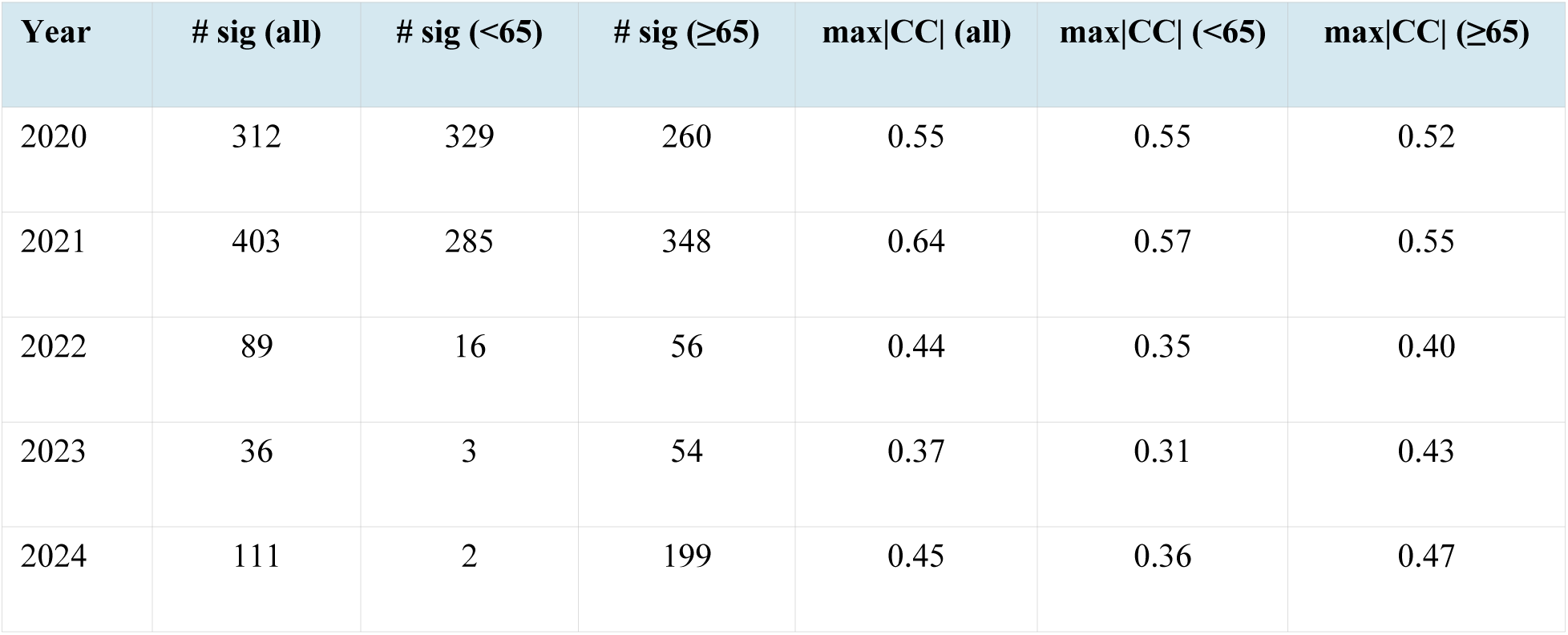
Age-band divergence over the pandemic. Counts of moderately associated variables (| CC| > 0.30, strict) and the single strongest |CC| for the all-age, under-65 (LT65) and 65– and – over (GE65) excess-death outcomes, by year (population-weighted, 2019-vintage predictors). The under-65 signal is confined to 2020–2021; the ≥65 signal persists and re-strengthens through 2024.

| Year | # sig (all) | # sig (<65) | # sig ($\geq 65$) | max$\vert CC\vert $ (all) | max$\vert CC\vert $ (<65) | max$\vert CC\vert $ ($\geq 65$) |
| --- | --- | --- | --- | --- | --- | --- |
| 2020 | 312 | 329 | 260 | 0.55 | 0.55 | 0.52 |
| 2021 | 403 | 285 | 348 | 0.64 | 0.57 | 0.55 |
| 2022 | 89 | 16 | 56 | 0.44 | 0.35 | 0.40 |
| 2023 | 36 | 3 | 54 | 0.37 | 0.31 | 0.43 |
| 2024 | 111 | 2 | 199 | 0.45 | 0.36 | 0.47 |

**Table S7.** The 11 super-clusters (Ward cut, k=11) and their size. Sizes count the 2,727 variables with a valid correlation (the 18 of the 2,745 screened predictors that lacked a usable cross-county correlation are omitted), matching the basis of Table 1.

| Super Cluster | Name | # clusters | # variables |
| --- | --- | --- | --- |
| 1 | Race, Ethnicity and Population by Age | 16 | 463 |
| 2 | Geography, Environment and Housing | 6 | 88 |
| 3 | Socioeconomic, Insurance and Demographic Characteristics | 14 | 412 |
| 4 | Vital Statistics and Total Physician Counts | 5 | 88 |
| 5 | Poverty, Insurance Coverage and Public Programs | 7 | 175 |
| 6 | Primary Care, Generalist and Diagnostic Physicians | 29 | 590 |
| 7 | Surgical, Procedural and Specialty Physicians | 17 | 266 |
| 8 | Commuting and Industry Employment | 2 | 48 |
| 9 | Aggregate Physician and Facility Totals | 10 | 180 |
| 10 | Hospital Telehealth and Advanced Imaging Services | 2 | 43 |
| 11 | Hospital Capacity and Nursing and Allied Staffing | 12 | 374 |

**Table S8.**
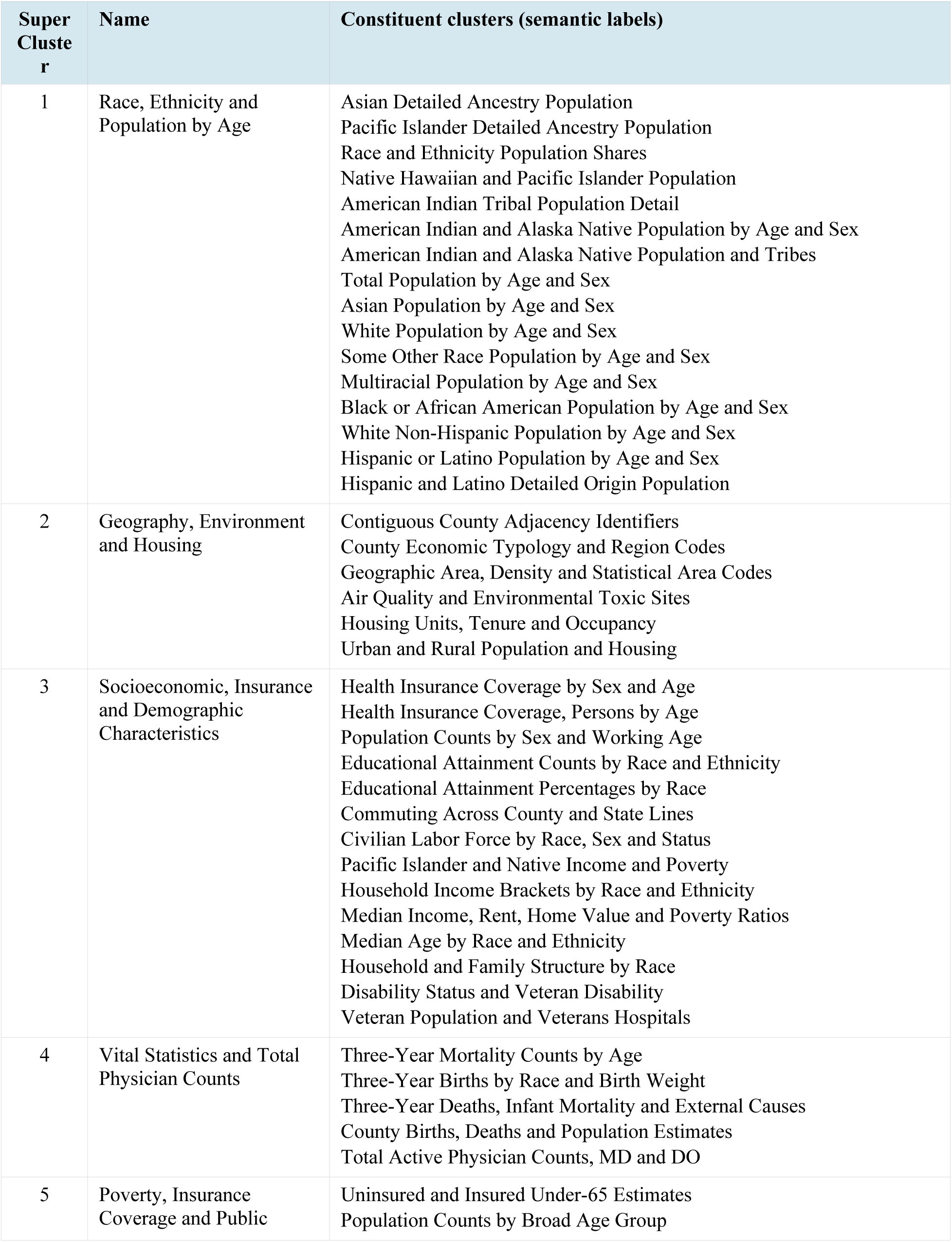
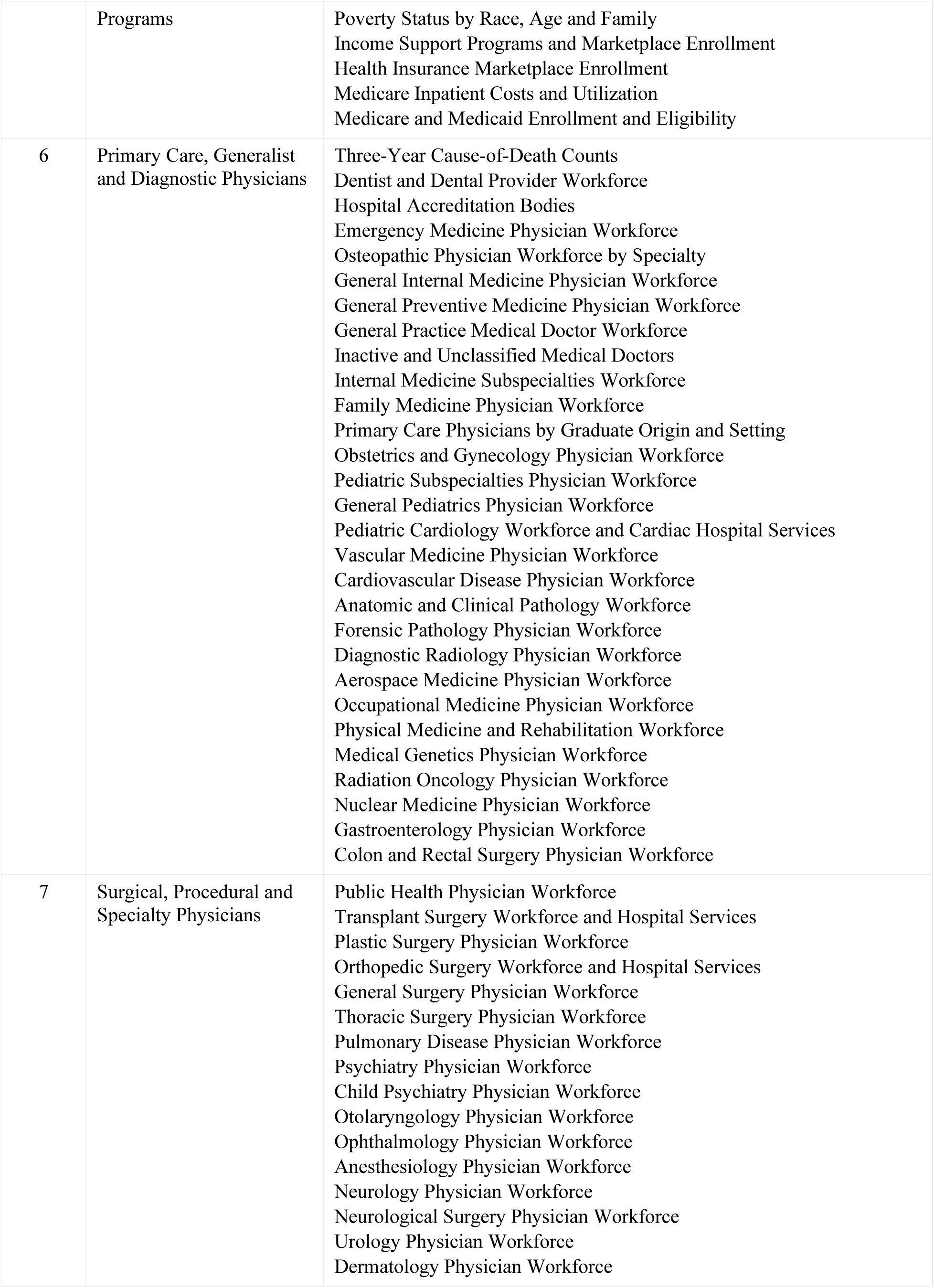
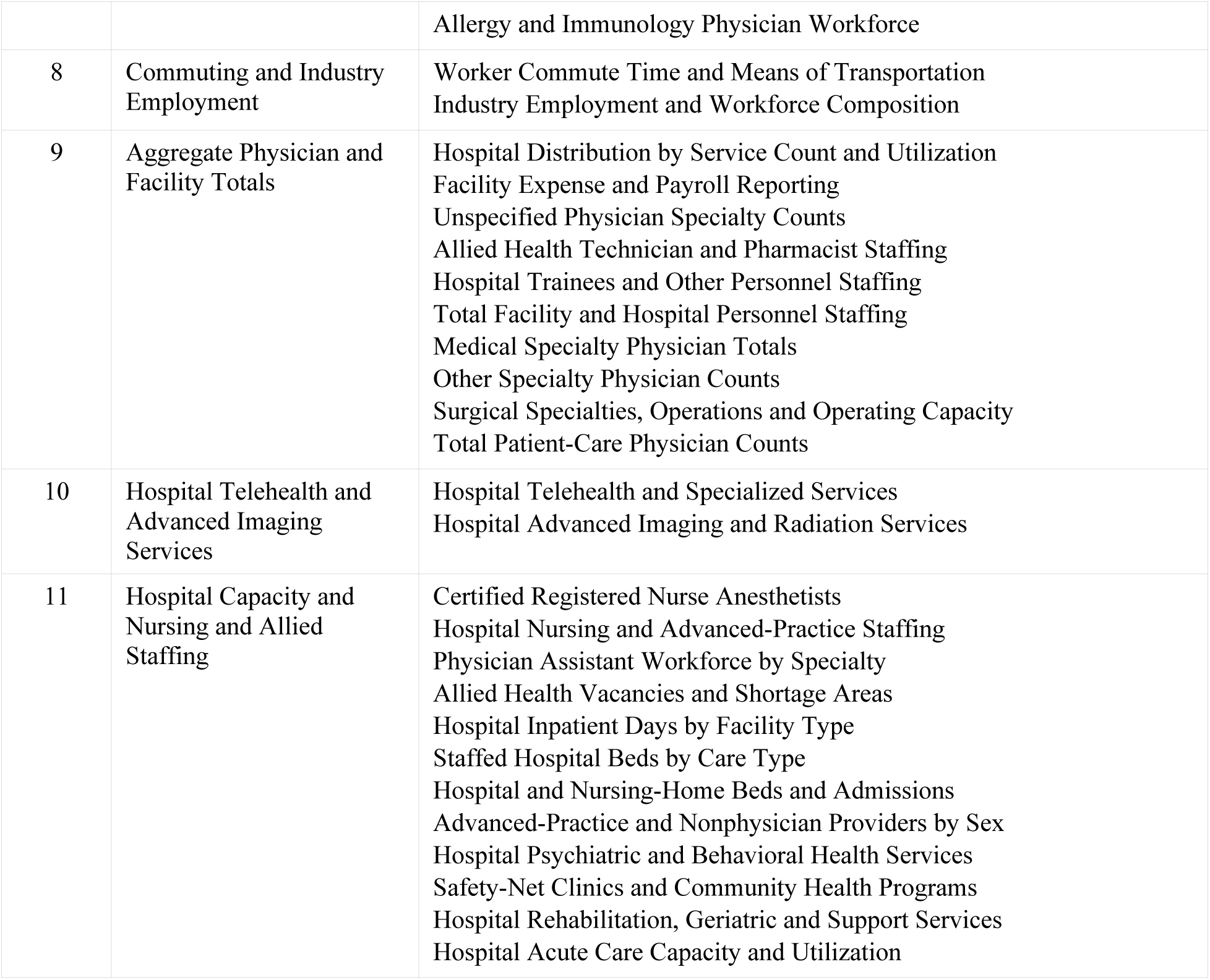
The 11 super-clusters and their constituent 120 semantic clusters (cluster labels).

**Table S9.** Top six moderately associated (|CC| > 0.30) distinct variables per super-cluster, with the signed CC at the year of maximum |CC| (Yr). Deduplicated: within each super-cluster only one representative — the highest-|CC| member — is shown per k=120 semantic cluster, so the listed variables are distinct measures rather than age/sex/race/vintage variants of the same one. The Collapsed column gives the number of other moderately associated variables in that cluster that the representative stands for; those variants are enumerated in Table_S9A_collapsed_variants.tsv (Supplementary Data). Where a super-cluster has fewer than six such clusters, fewer than six rows appear. Same basis and super-cluster ordering as Table 1; SC10 (Hospital Telehealth and Advanced Imaging Services) has no variable reaching |CC| > 0.30. Variable names are AHRF descriptions (verbatim, including source spellings).

| SC | Super-cluster (# variables) | # | Representative variable | CC | Yr | Collapsed |
| --- | --- | --- | --- | --- | --- | --- |
| 3 | Socioeconomic, Insurance and Demographic Characteristics (412) | 1 | Per 25+ with 4+ Year College White non Hispanic | -0.62 | 2021 | 21 |
| | | 2 | #White non Hispanic Household Income \$100 000+ | -0.61 | 2021 | 25 |
|  |  | 3 | % Female 40 64 without Health Insurance | +0.61 | 2021 | 35 |
|  |  | 4 | Personal Persons 18 64 with Health Insurance | -0.60 | 2021 | 13 |
|  |  | 5 | Personal Persons 25+ with 4+ Years College White | -0.59 | 2021 | 11 |
|  |  | 6 | Ratio of Income to Poverty Level 1.25 1.49 | +0.59 | 2021 | 15 |
| 5 | Poverty, Insurance Coverage and Public Programs (175) | 1 | Persons <65 <= 400% of Poverty | +0.64 | 2021 | 11 |
|  |  | 2 | # <19 with Health Insurance <=200% Poverty | +0.62 | 2021 | 29 |
|  |  | 3 | Persons Age 0 17 in Poverty | +0.59 | 2021 | 25 |
|  |  | 4 | Inpatient Per Capita Actual Medicare Cost | +0.55 | 2020 | 2 |
|  |  | 5 | Medicare Fee for Service Beneficiaries Average Hierarchical Condition Categories Score | +0.54 | 2020 | 9 |
|  |  | 6 | Food Stamp Supplemental Nutrition Assistance Program Recipients | +0.42 | 2021 | 0 |
| 8 | Commuting and Industry Employment (48) | 1 | # 16+ Workers Public Transplant | +0.55 | 2020 | 10 |
|  |  | 2 | % Population Did Not Work 16 64 | +0.54 | 2021 | 5 |
| 1 | Race, Ethnicity and Population by Age (463) | 1 | Hispanic Latino Origin Ecuadorian Population | +0.55 | 2020 | 20 |
|  |  | 2 | Population Black African Hispanic Latino Female | +0.53 | 2020 | 8 |
|  |  | 3 | American Indian South American Population | +0.51 | 2020 | 2 |
|  |  | 4 | Population Some Other Race Female 65 74 | +0.50 | 2020 | 27 |
|  |  | 5 | Population White Non Hispanic Female 55 59 | -0.49 | 2020 | 29 |
|  |  | 6 | Asian Bangladeshi Population | +0.48 | 2020 | 2 |
| 2 | Geography, Environment and Housing (88) | 1 | # Owner Occupied Housing Units | -0.49 | 2020 | 3 |
|  |  | 2 | Population Density per Square Mile | +0.49 | 2020 | 1 |
|  |  | 3 | Low Education Typology Code | +0.41 | 2021 | 8 |
|  |  | 4 | Census Urbanized Areas Housing | -0.33 | 2024 | 10 |
| 9 | Aggregate Physician and Facility Totals (180) | 1 | Medical Specialty Total 45 54 | -0.39 | 2021 | 10 |
|  |  | 2 | M.D.'s Total Other | -0.39 | 2021 | 3 |
|  |  | 3 | Other Specialty Total 55 64 | -0.39 | 2021 | 9 |
|  |  | 4 | Surgical Specialty Total 45 54 | -0.34 | 2021 | 1 |
|  |  | 5 | All Other Personnel Partial Time | -0.31 | 2021 | 0 |
| 4 | Vital Statistics and Total Physician Counts (88) | 1 | Number Employed 16+ | -0.50 | 2021 | 2 |
|  |  | 2 | Total M.D.'s 55 64 | -0.40 | 2021 | 9 |
|  |  | 3 | 3 Year Total Births | +0.34 | 2020 | 0 |
| 6 | Primary Care, Generalist and Diagnostic Physicians (590) | 1 | Medical Doctors Inactive Female | -0.46 | 2021 | 5 |
|  |  | 2 | # Active Dentists 55 64 | -0.42 | 2021 | 11 |
|  |  | 3 | Physician Primary Care Patient Care | -0.42 | 2021 | 10 |
|  |  | 4 | General Internal Medical Medicine 55 64 | -0.40 | 2021 | 8 |
|  |  | 5 | Pediatrics General 45 54 | -0.38 | 2021 | 5 |
|  |  | 6 | Emergency Medical Medicine Patient Care<br>Hospital Full Time Staff | -0.35 | 2021 | 3 |
| 7 | Surgical,<br>Procedural and<br>Specialty<br>Physicians (266) | 1 | Psychiatry 55 64 | -0.38 | 2021 | 6 |
|  |  | 2 | Child Psychiatric Total Patient Care | -0.36 | 2021 | 6 |
|  |  | 3 | Dermatology Patient Care Office Based | -0.34 | 2021 | 4 |
|  |  | 4 | Ophthalmology Patient Care Office Based | -0.33 | 2021 | 4 |
|  |  | 5 | Allergy & Immunology Total | -0.33 | 2021 | 2 |
|  |  | 6 | Anesthesiology 55 64 | -0.33 | 2021 | 4 |
| 11 | Hospital Capacity<br>and Nursing and<br>Allied Staffing<br>(374) | 1 | # Education Healthcare Social Assist Work | -0.42 | 2021 | 1 |
|  |  | 2 | Chiropractors with National Provider Identifier | -0.37 | 2021 | 7 |
|  |  | 3 | # Hospices | +0.30 | 2021 | 0 |
| 10 | Hospital Telehealth<br>and Advanced<br>Imaging Services<br>(43) | - | (none reaching the moderate band) | - | - | - |

**Table S10.** Excess death by population-weighted quintile of the advantage index. The index is the mean of the 200 socioeconomic and demographic variables reaching |CC| > 0.45, each standardized across counties and sign-aligned so that a high score corresponds to lower excess death. Quintiles are cut on population, so each holds about 65 million people and the county counts differ. Excess-death figures are population-weighted means of the proportion above each county own pre-pandemic expectation. The rural population share is from the 2020 Census; metropolitan status is rural-urban continuum code 1 to 3. The index correlates with excess death at −0.55 in 2020 and −0.56 in 2021, falling to −0.04 by 2024. The gradient is monotonic in 2020 and 2021, in 2022 to 2024 and in the non-metropolitan share; the 2.0-fold contrast of 2020 and 2021 narrows to 1.7-fold in 2022 to 2024, on a level roughly five times lower. Taken year by year, the most disadvantaged fifth had 0.285 in 2020 and 0.264 in 2021 against 0.152 and 0.121 in the most advantaged.

| Quintile of the advantage index | Counties | Population (millions) | Excess death 2020 and 2021 | Excess death 2022 to 2024 | Rural population (%) | Non-metropolitan counties (%) |
| --- | --- | --- | --- | --- | --- | --- |
| Q1 most disadvantaged | 462 | 65.8 | 0.274 | 0.050 | 13.0 | 74.9 |
| Q2 | 587 | 64.6 | 0.211 | 0.045 | 19.7 | 64.4 |
| Q3 | 629 | 66.0 | 0.177 | 0.044 | 22.2 | 58.0 |
| Q4 | 561 | 66.8 | 0.159 | 0.041 | 21.2 | 54.2 |
| Q5 most advantaged | 539 | 65.8 | 0.136 | 0.030 | 22.7 | 44.7 |

**Table S11.** Excess death and the advantage index by Census region and division. Region and division are the AHRF code variables already in the predictor set. Excess death is the population-weighted mean proportion for 2020 and 2021; Q1 is the most disadvantaged population-weighted quintile of Table S10. Division differences are larger than region differences, because each region mixes advantaged and disadvantaged divisions. The extremes are West South Central, at 0.241 with 44.3% of its population in the most disadvantaged quintile, and New England, at 0.136 with none. Metropolitan counties, 1,144 of them holding 284.0 million people, had 0.190; non-metropolitan counties, 1,634 holding 45.0 million, had 0.200.

| Census region or division | Counties | Population (millions) | Excess death 2020 and 2021 | Mean advantage index | Population in Q1 (%) |
| --- | --- | --- | --- | --- | --- |
| Northeast | 216 | 57.6 | 0.192 | +0.20 | 18.2 |
| Midwest | 863 | 68.2 | 0.173 | +0.37 | 0.7 |
| South | 1,344 | 125.2 | 0.202 | −0.27 | 23.7 |
| West | 355 | 77.9 | 0.190 | −0.24 | 32.3 |
| New England | 67 | 15.1 | 0.136 | +0.67 | 0.0 |
| Middle Atlantic | 149 | 42.5 | 0.211 | +0.04 | 24.6 |
| East North Central | 430 | 47.3 | 0.181 | +0.31 | 0.2 |
| West North Central | 433 | 20.9 | 0.153 | +0.51 | 2.0 |
| South Atlantic | 574 | 65.6 | 0.175 | −0.11 | 16.2 |
| East South Central | 361 | 19.3 | 0.210 | −0.02 | 6.1 |
| West South Central | 409 | 40.3 | 0.241 | −0.65 | 44.3 |
| Mountain | 212 | 24.4 | 0.200 | −0.12 | 19.9 |
| Pacific | 143 | 53.5 | 0.186 | −0.30 | 37.9 |

**Table S12.** Cross-vintage agreement on moderate-association status (|CC| > 0.30), restricted to the variables whose vintage value actually changed (775 backward and 779 forward base-matched variables that are identical by construction — not rolled back or not refreshed — are excluded as uninformative). The 2015 comparison is a retrospective cutoff of the 2019–2020 release, not an independent download; agreement is still partly carried by concordant sub-threshold variables.

| Comparison | Refreshed<br>variables | Moderate in<br>both | Neither in<br>both | Agreement |
| --- | --- | --- | --- | --- |
| 2015 vs 2019–2020 | 1,617 | 384 | 1,192 | 97.5% |
| 2019–2020 vs 2023–2024 | 1,971 | 364 | 1,548 | 97.0% |

**Table S13.** Sensitivity of the variable–death correlations to the county weighting scheme. Each county is weighted by its 2019 population raised to the power p (p = 0 unweighted; p = 1.0 the primary full-population analysis). For each scheme: the Kish effective sample size; the percentage of the 2,727 analyzed variables whose maximum |CC| over 2020–2024 reaches the moderate (> 0.30) and strong (> 0.45) bands; the median of that maximum |CC|; and the single largest |CC|. Poverty-share, uninsurance and college-attainment measures lead under every scheme, in varying order, and the magnitudes scale with p.

| Weight power $p$ | County weighting | N_eff | % $\vert CC\vert > 0.30$ | % $\vert CC\vert > 0.45$ | Median max $\vert CC\vert $ | Max $\vert CC\vert $ |
| --- | --- | --- | --- | --- | --- | --- |
| 0 | Unweighted | 2,778 | 5.6% | 0% | 0.082 | 0.436 |
| 0.5 | Square-root of population | 1,512 | 11.8% | 2.6% | 0.130 | 0.524 |
| 0.75 | Population<sup>0.75</sup> | 695 | 18.1% | 4.4% | 0.160 | 0.586 |
| 1.0 | Full population (primary) | 279 | 24.1% | 7.4% | 0.181 | 0.642 |

**Table S14.** What is and is not robust in these analyses. Direction, ranking and the contrast between the two families are stable across every sensitivity analysis we ran. The magnitude of the correlations and the proportion crossing a band are not: both scale with the weight given to large counties. Claims of robustness in this paper refer to the first group.

| Aspect of robustness | Holds? | Evidence |
| --- | --- | --- |
| Direction and sign | Yes | Three sign reversals backward and two forward among the 397 and 399 variables reaching $\vert CC\vert > 0.30$ (Figure S6) |
| Ranking of the leading correlates | Yes | Poverty-share, uninsurance and college-attainment measures lead at every weighting and under Spearman, in varying order |
| Magnitude of the correlations | No | Largest $\vert CC\vert $ 0.64, 0.59, 0.52, 0.44 at weight powers $p = 1.0, 0.75, 0.5, 0$ (Table S13) |
| Threshold classification | No | Share reaching $\vert CC\vert > 0.30$ is 24.1%, 18.1%, 11.8%, 5.6% across the same weightings (Table S13) |
| Socioeconomic versus health-system contrast | Yes | Holds at every weighting, under Spearman, and under the data-driven two-way partition |

**Table S15.** Sensitivity of the variable–death correlations to the choice of correlation coefficient, by super-cluster. For each of the 11 semantic super-clusters: the number of variables (N) and, under population-weighted Pearson (P) and population-weighted Spearman (S) correlation, the percentage of variables whose maximum |CC| over the all-age years 2020–2024 reaches the moderate band (> 0.30), the number reaching the strong band (> 0.45), and the single largest |CC| (2,727 variables with a valid CC, as in Table 1). Spearman gives a larger moderate share in every super-cluster but fewer strong-band variables (168 against 202), and the qualitative pattern is preserved: the socioeconomic and demographic super-clusters dominate, accounting for 165 of the 168 strong-band variables under Spearman; among the health system super-clusters two contain one strong-band variable each under Pearson (super-clusters 4 and 6) and three variables reach it under Spearman (one each in super-clusters 4, 6 and 11). Spearman coefficients are population-weighted rank correlations, computed as weighted empirical-distribution ranks followed by a weighted Pearson correlation, the same method used for Table S16.

| Super-cluster | N | % mod<br>(P) | % mod<br>(S) | strong n<br>(P) | strong n<br>(S) | max \|<br>CC\|<br>(P) | max \|<br>CC\|<br>(S) |
| --- | --- | --- | --- | --- | --- | --- | --- |
| 1. Race, Ethnicity and Population by Age | 463 | 32.4 | 49.5 | 26 | 11 | 0.55 | 0.49 |
| 2. Geography, Environment and Housing | 88 | 29.5 | 35.2 | 3 | 1 | 0.49 | 0.45 |
| 3. Socioeconomic, Insurance and Demographic Characteristics | 412 | 53.4 | 60.7 | 115 | 98 | 0.62 | 0.66 |
| 4. Vital Statistics and Total Physician Counts | 88 | 15.9 | 46.6 | 1 | 1 | 0.50 | 0.50 |
| 5. Poverty, Insurance Coverage and Public Programs | 175 | 46.9 | 54.9 | 49 | 49 | 0.64 | 0.67 |
| 6. Primary Care, Generalist and Diagnostic Physicians | 590 | 12.7 | 19.2 | 1 | 1 | 0.46 | 0.48 |
| 7. Surgical, Procedural and Specialty Physicians | 266 | 12.0 | 13.9 | 0 | 0 | 0.38 | 0.40 |
| 8. Commuting and Industry Employment | 48 | 35.4 | 43.8 | 7 | 6 | 0.55 | 0.57 |
| 9. Aggregate Physician and Facility Totals | 180 | 15.6 | 22.2 | 0 | 0 | 0.39 | 0.43 |
| 10. Hospital Telehealth and Advanced Imaging Services | 43 | 0.0 | 4.7 | 0 | 0 | 0.29 | 0.35 |
| 11. Hospital Capacity and Nursing and Allied Staffing | 374 | 3.2 | 5.3 | 0 | 1 | 0.42 | 0.47 |

**Table S16.** The twenty strongest correlations, with the matching Spearman coefficient. SC is the semantic super-cluster; Year is the year of maximum |CC|; Pearson is the population-weighted Pearson correlation reported throughout the paper; Spearman is the population-weighted rank correlation, computed as weighted empirical-distribution ranks followed by a weighted Pearson correlation. Sign agrees for all twenty, and the Spearman coefficient is larger in magnitude for all twenty, which is consistent with monotonic but not exactly linear relationships. All twenty lie in the socioeconomic super-clusters 3 and 5.

| SC | Variable | Year | Pearson | Spearman |
| --- | --- | --- | --- | --- |
| 5 | Persons under 65 at or below 400% of poverty | 2021 | +0.64 | +0.67 |
| 5 | Persons under 19 at or below 200% of poverty | 2021 | +0.64 | +0.66 |
| 5 | Persons under 65 at or below 200% of poverty | 2021 | +0.63 | +0.65 |
| 5 | Persons under 19 at or below 400% of poverty | 2021 | +0.63 | +0.65 |
| 5 | Persons under 19 at or below 138% of poverty | 2021 | +0.62 | +0.65 |
| 5 | Persons under 19 with health insurance, at or below 200% of poverty | 2021 | +0.62 | +0.64 |
| 3 | Adults 25+ with 4+ years college, White non-Hispanic (%) | 2021 | -0.62 | -0.64 |
| 3 | White non-Hispanic households with income \$100,000+ | 2021 | -0.61 | -0.64 |
| 5 | Persons under 19 with health insurance, at or below 400% of poverty | 2021 | +0.61 | +0.64 |
| 3 | Females 40–64 without health insurance (%) | 2021 | +0.61 | +0.66 |
| 3 | Females 18–64 without health insurance (%) | 2021 | +0.61 | +0.65 |
| 3 | Nonveterans 25+ with high school diploma or more | 2021 | -0.60 | -0.64 |
| 5 | Persons 40–64 without health insurance (%) | 2021 | +0.60 | +0.65 |
| 3 | Persons 18–64 with health insurance | 2021 | -0.60 | -0.62 |
| 5 | Persons under 19 with health insurance, at or below 138% of poverty | 2021 | +0.60 | +0.62 |
| 5 | Persons 18–64 without health insurance (%) | 2021 | +0.60 | +0.63 |
| 5 | Persons 18–64 at or below 400% of poverty | 2021 | +0.60 | +0.63 |
| 3 | Persons 25+ with 4+ years college, White | 2021 | -0.59 | -0.61 |
| 3 | Females under 65 without health insurance (%) | 2021 | +0.59 | +0.64 |
| 3 | Persons with income 1.25–1.49 times the poverty level | 2021 | +0.59 | +0.63 |

### Supplementary Figures

**Figure S1.**
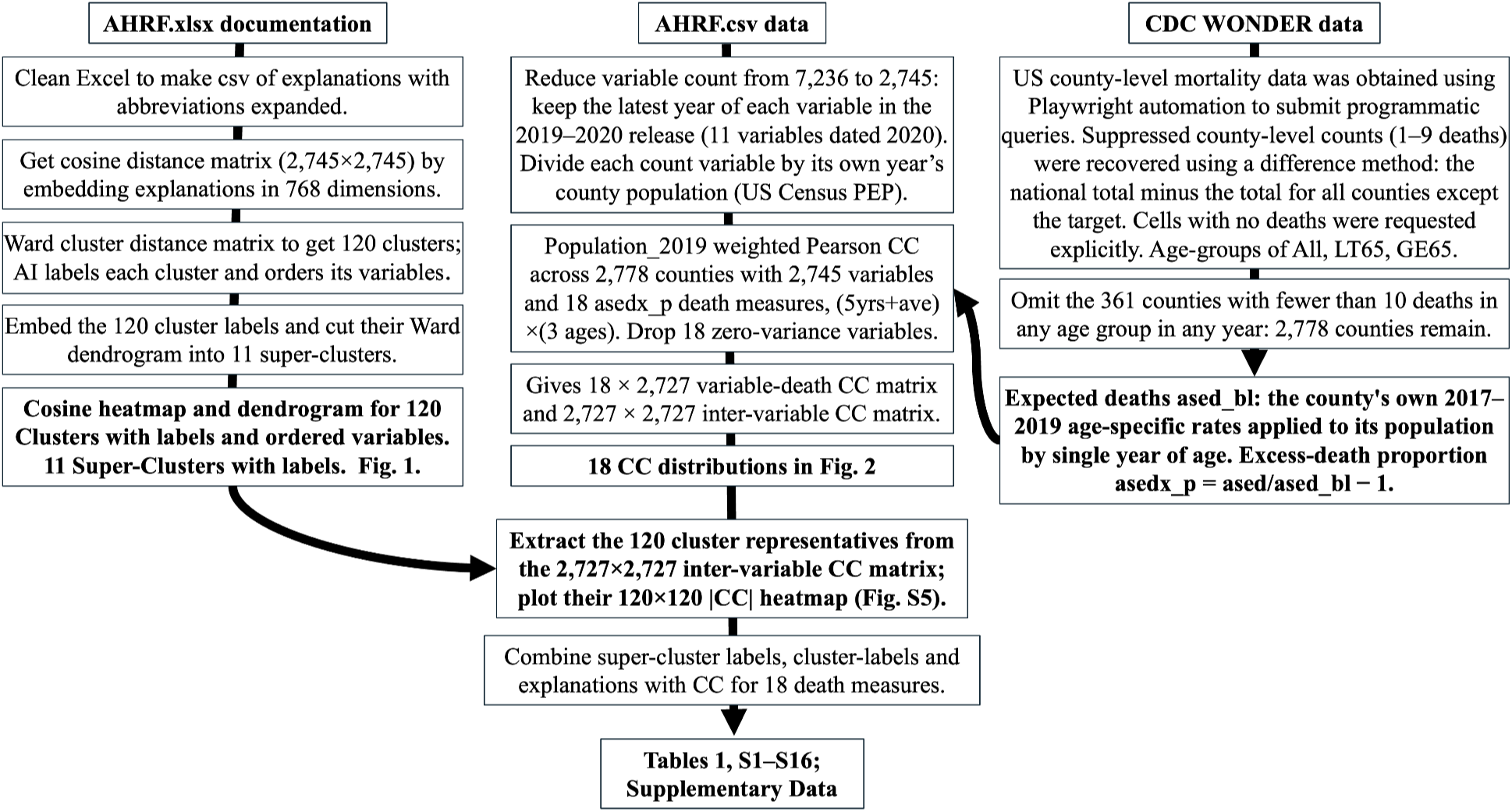
Pipeline overview, from source files (AHRF technical documentation, AHRF data, CDC WONDER mortality) to the analysis variable set and the results tables. Documents the reduction from 7,236 raw AHRF variable columns (all vintages) to the 2,745 described variables used downstream.

**Figure S2.**
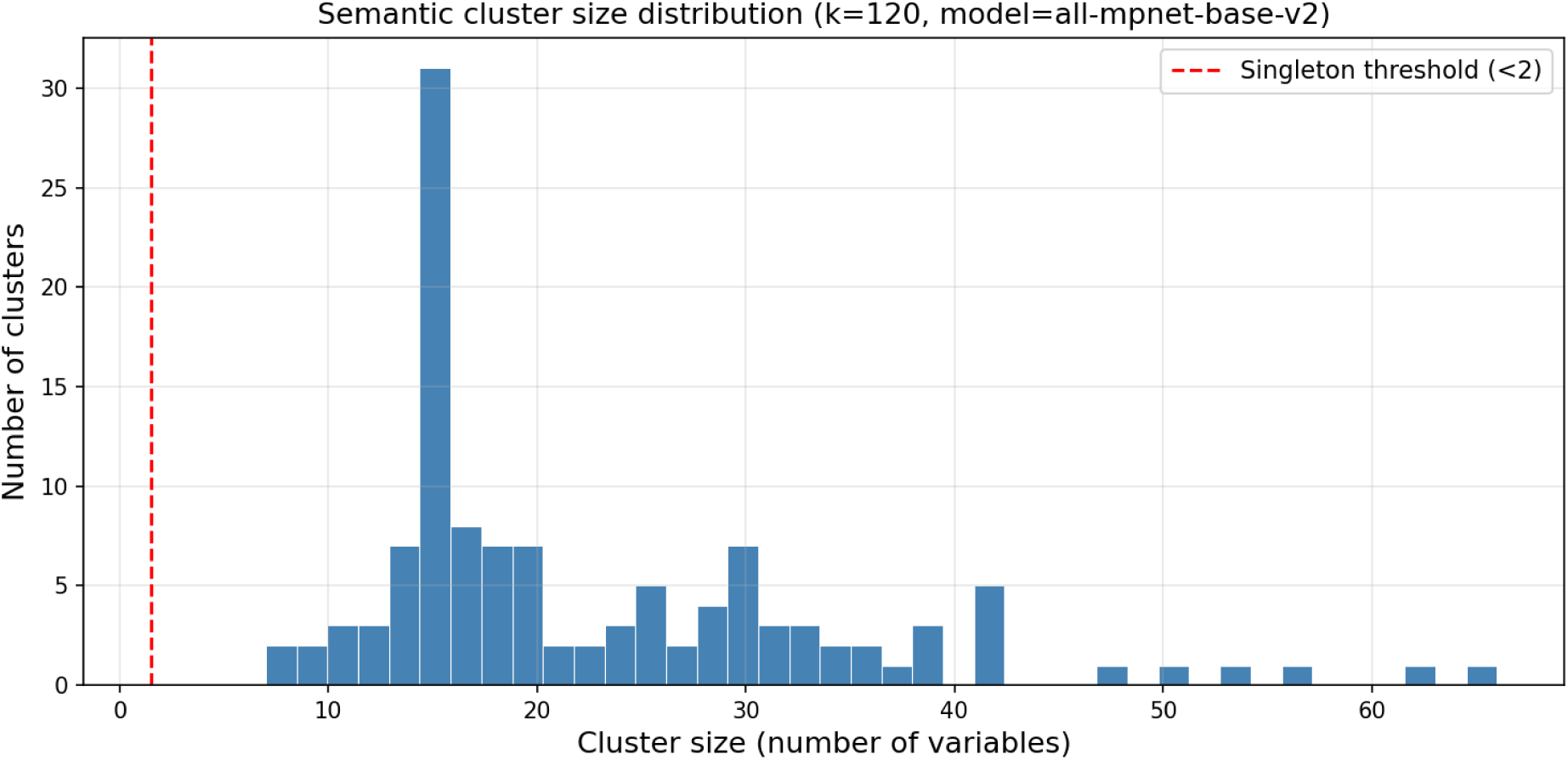
Distribution of variable counts across the 120 semantic clusters (sizes 7–66 variables, median 18).

**Figure S3.**
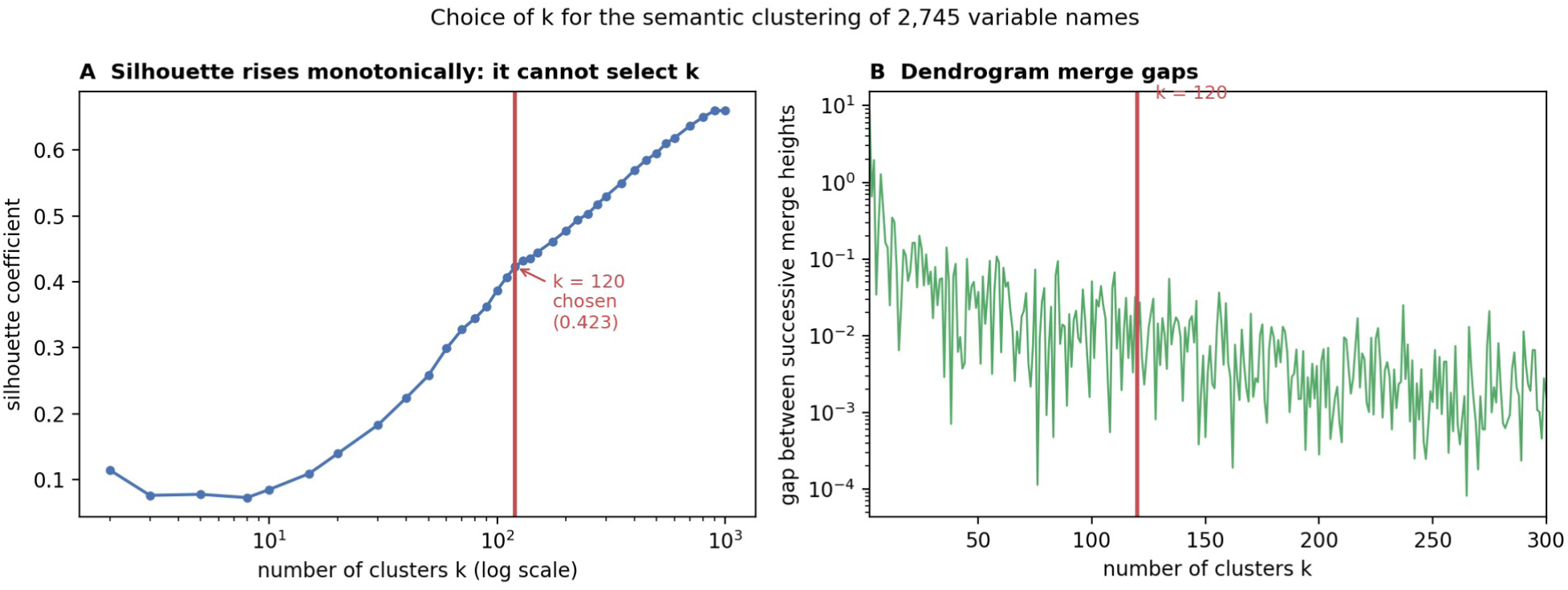
Silhouette coefficient against the number of clusters k, for the cleaned-name sentence-transformer embeddings of the 2,745 variable names. (A) The silhouette rises monotonically from k = 2 to k = 1000 and never peaks, so it cannot identify an optimal k; the value at the chosen k = 120 is 0.423. (B) Gaps between successive dendrogram merge heights, log scale: the largest gaps fall at k = 2, 4 and 7, reflecting the top-level split between the demographic and health-system halves, with nothing distinguishing k = 120 from its neighbours. k = 120 is a granularity choice and not an optimum.

**Figure S4.**
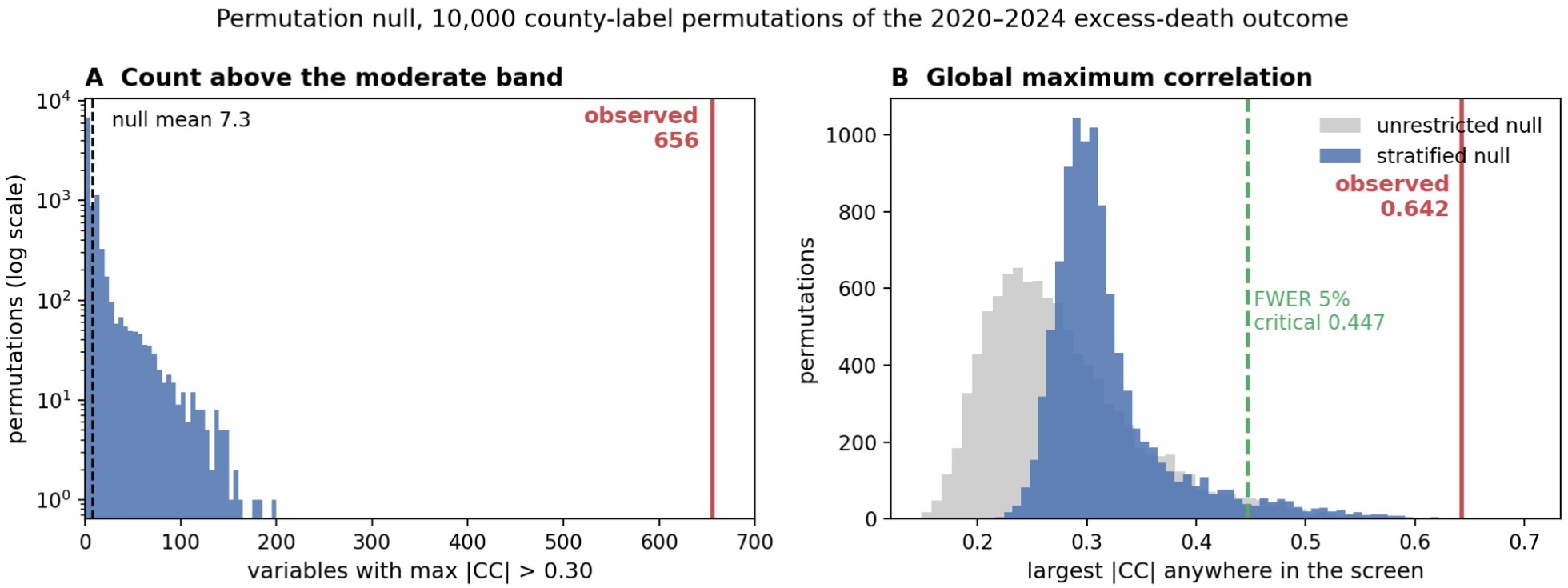
Permutation null for the variable screen. County labels of the whole five-year excess-death block were permuted together, holding the predictors and the county weights at their own rows, and the maximum absolute correlation coefficient over 2020–2024 was recomputed for all 2,727 variables; 10,000 permutations. The primary null is stratified by 2019 county population in 20 equal-count strata, so an outcome value can move only to a county of similar size. (A) Number of variables reaching |CC| > 0.30 in each permutation, log scale: the null mean is 7.3 and no permutation of the 10,000 reaches the observed 656. (B) Largest |CC| anywhere in the screen, for the stratified null (blue) and the unrestricted null (grey). The observed 0.642 is marked in red; the dashed green line at 0.447 is the 95th percentile of the stratified null, the critical value controlling the family-wise error rate at 5% across all 13,725 tests.

**Figure S5.**
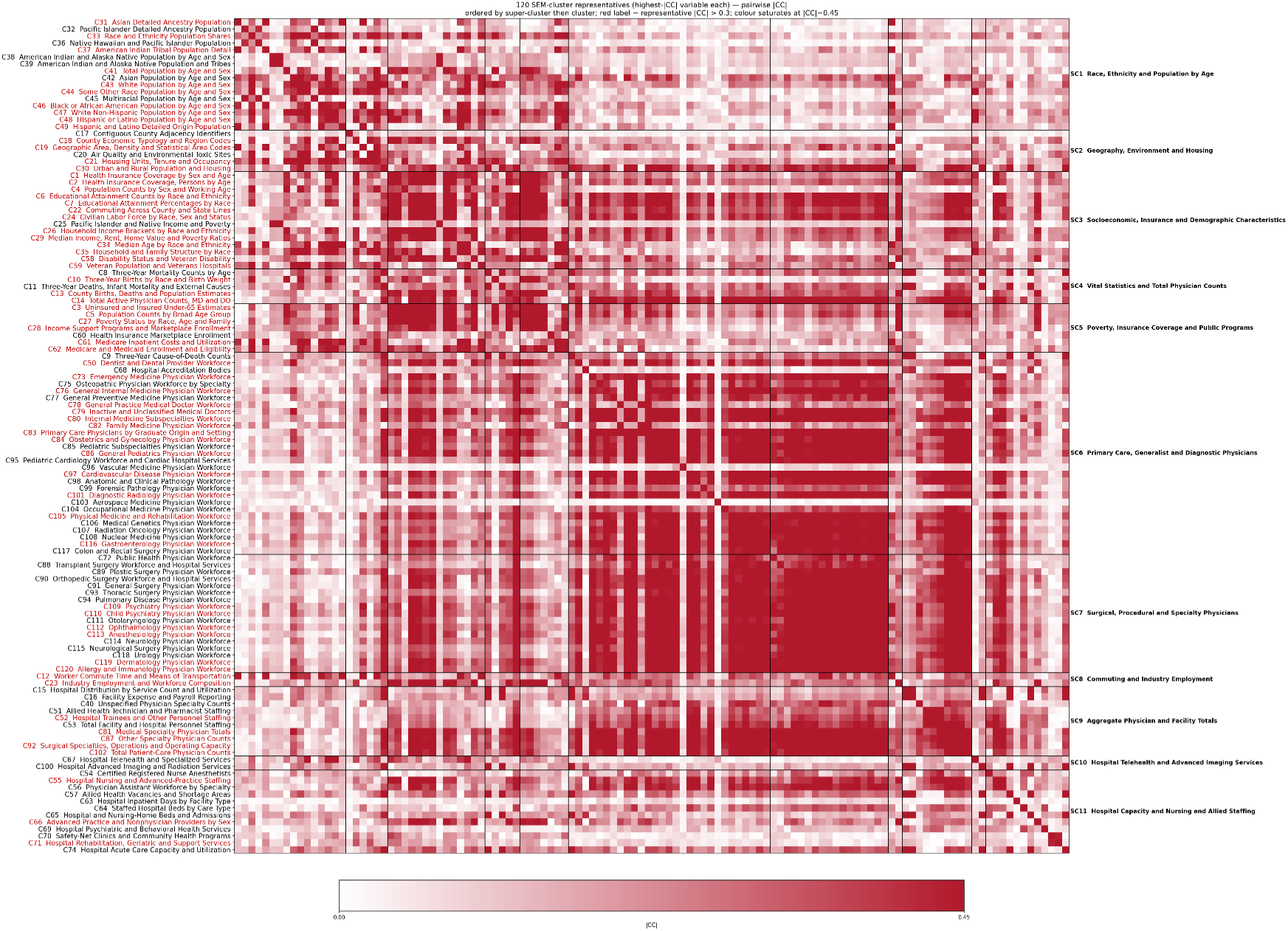
120×120 representative-variable |CC| heatmap (each cluster’s highest-|CC| member); cell shading is |CC| magnitude (white→red, capped at 0.45). A cluster label is printed in red when its representative variable reaches |CC| > 0.30 (i.e. the cluster’s strongest variable is moderately associated) — a looser criterion than Figure 1, which reddens a cluster only when ≥3 of its variables reach the moderate band in ≥2 years. The bright blocks are the demographic and socioeconomic super-clusters.

**Figure S6.**
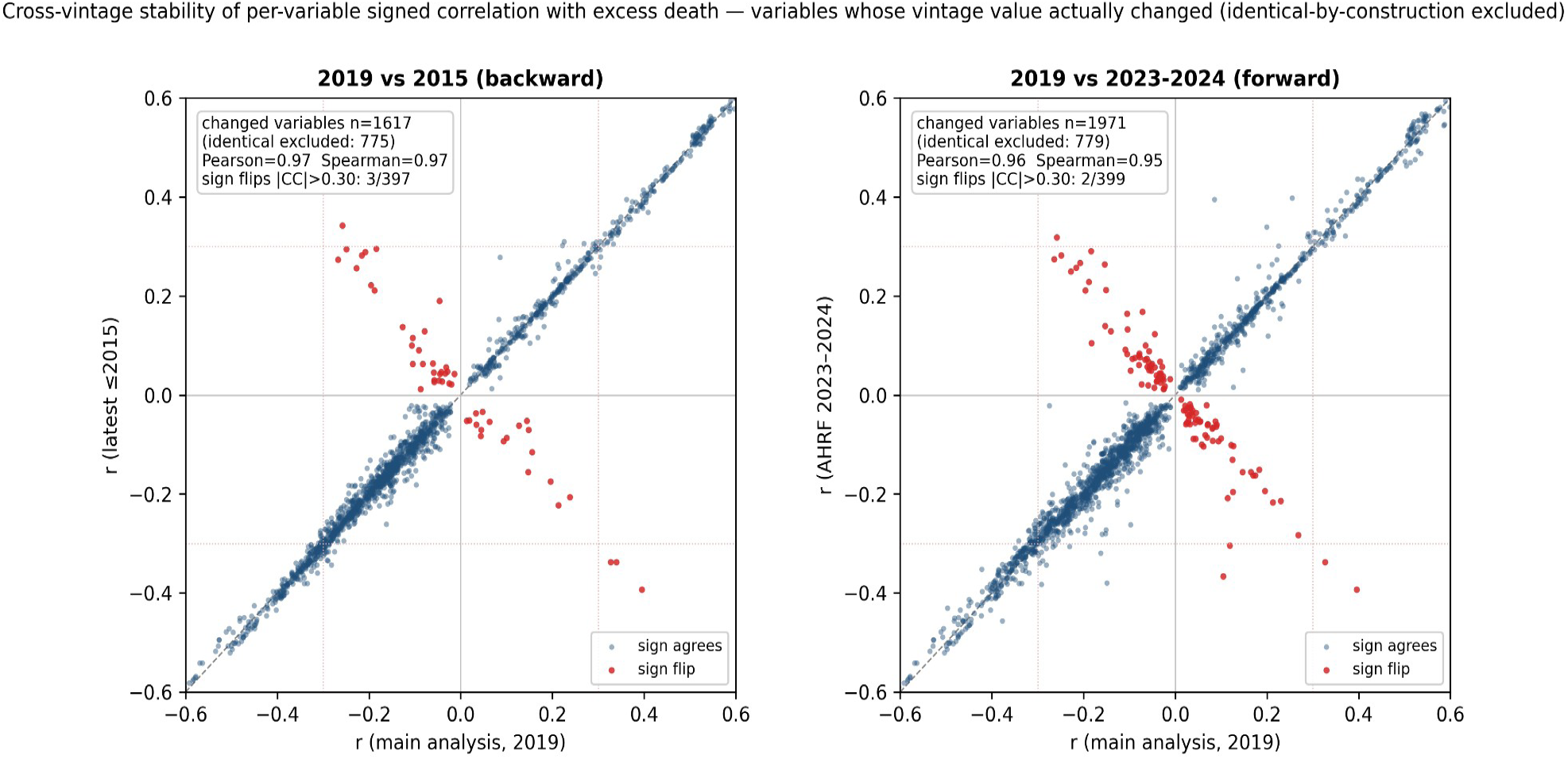
Cross-vintage stability of the per-variable correlation with excess death. The comparison is restricted to the variables whose vintage value actually changed between releases: variables that are identical by construction — not rolled back (≤2015) or not refreshed (2023–2024) — are excluded as uninformative (775 excluded backward, leaving 1,617 tested; 779 excluded forward, leaving 1,971 tested). Each plotted point is one such variable; the axes are its signed correlation coefficient (the per-variable maximum |CC| over 2020–2024, with sign) in the main (2019–2020) analysis (x) versus the backward (latest year ≤ 2015; left panel) and forward (AHRF 2023–2024 release; right panel) sensitivity analyses (y). The dashed line is identity; dotted lines mark |CC| = 0.30. Points are red where the sign disagrees between vintages; among the variables reaching |CC| > 0.30 in the main analysis three reverse sign backward and two do forward. Each panel annotates the Pearson and Spearman coefficients, the number of refreshed variables tested, and the number of identical-by-construction variables excluded.

**Figure S7.**
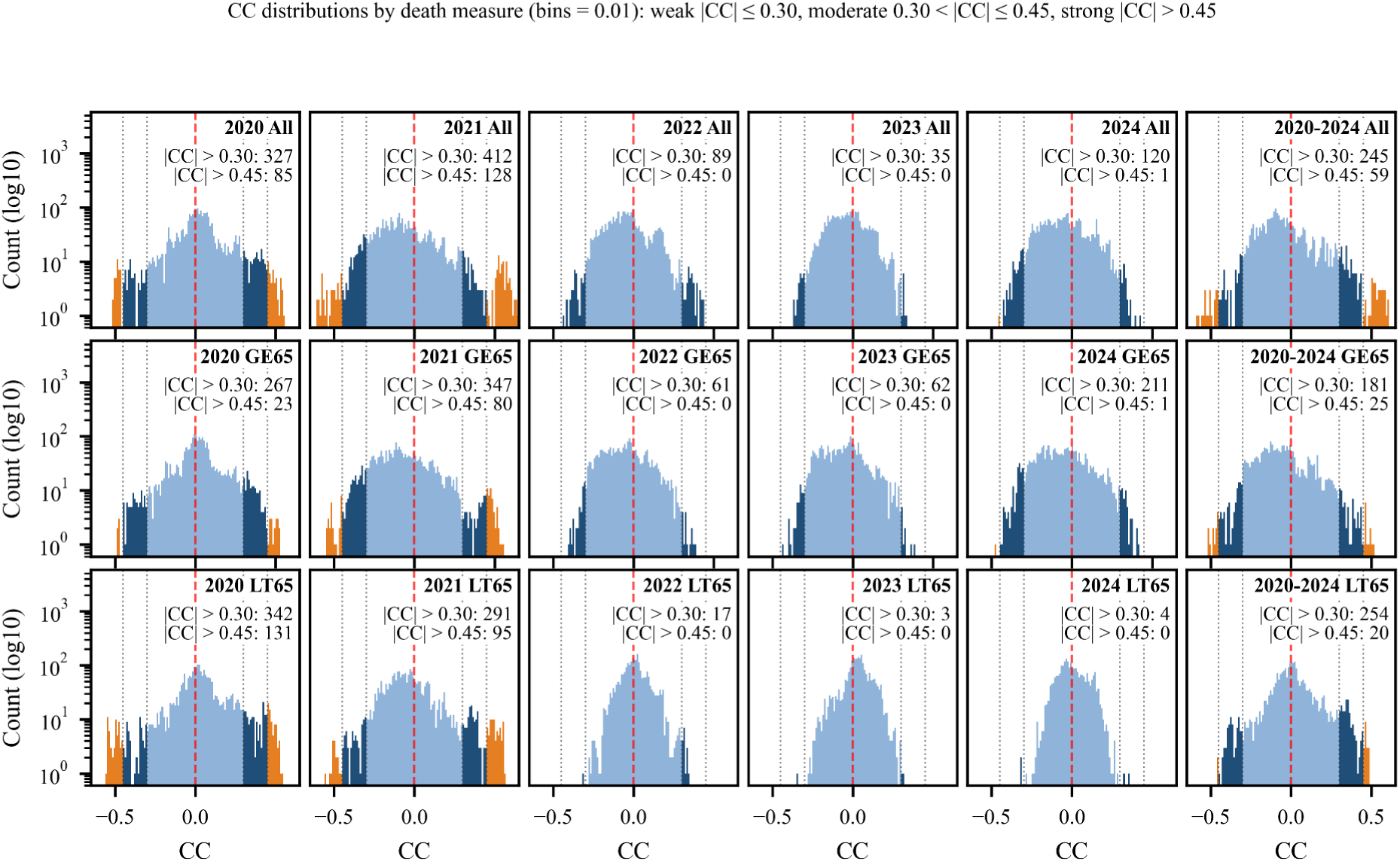
Variable–death CC distributions for the 2015 backward-cutoff vintage (population-weighted). Columns: death years 2020–2024 plus pooled 2020–2024; rows: All / Ages ≥65 / Ages <65. log₁₀ count on the y-axis; dark blue = moderate (|CC| > *0.30), orange = strong (|CC| > 0.45); red line at CC = 0*.

**Figure S8.**
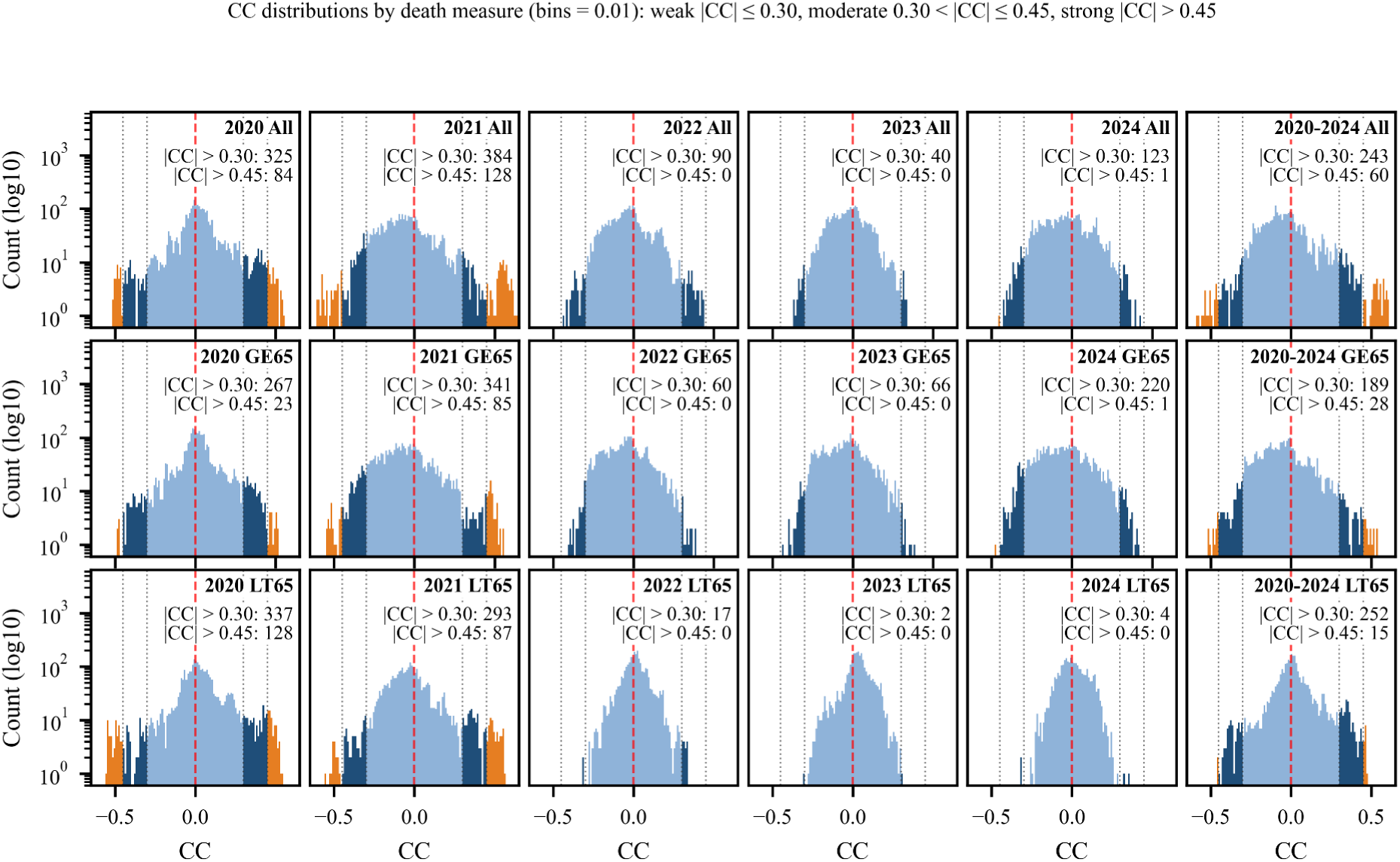
Variable–death CC distributions for the AHRF 2023–2024 vintage, same axes and coloring as Figure S7. The moderate and strong tails are populated comparably to Figure S7 (2015) and to the 2019 standard run — the visual counterpart of the high variable-level vintage agreement (Table S12; Figure S6).

